# Gastrointestinal involvement attenuates COVID-19 severity and mortality

**DOI:** 10.1101/2020.09.07.20187666

**Authors:** Alexandra E. Livanos, Divya Jha, Francesca Cossarini, Ana S. Gonzalez-Reiche, Minami Tokuyama, Teresa Aydillo, Tommaso L. Parigi, Irene Ramos, Katie Dunleavy, Brian Lee, Rebekah Dixon, Steven T. Chen, Gustavo Martinez-Delgado, Satish Nagula, Huaibin M. Ko, Benjamin S. Glicksberg, Girish Nadkarni, Elisabet Pujadas, Jason Reidy, Steven Naymagon, Ari Grinspan, Jawad Ahmad, Michael Tankelevich, Ronald Gordon, Keshav Sharma, Jane Houldsworth, Graham J. Britton, Alice Chen-Liaw, Matthew P. Spindler, Tamar Plitt, Pei Wang, Andrea Cerutti, Jeremiah J. Faith, Jean-Frederic Colombel, Ephraim Kenigsberg, Carmen Argmann, Miriam Merad, Sacha Gnjatic, Noam Harpaz, Silvio Danese, Carlos Cordon-Cardo, Adeeb Rahman, Nikhil A. Kumta, Alessio Aghemo, Francesca Petralia, Harm van Bakel, Adolfo Garcia-Sastre, Saurabh Mehandru

## Abstract

Given that gastrointestinal (GI) symptoms are a prominent extrapulmonary manifestation of coronavirus disease 2019 (COVID-19), we investigated intestinal infection with severe acute respiratory syndrome coronavirus-2 (SARS-CoV-2) and its effect on disease pathogenesis. SARS-CoV-2 was detected in small intestinal enterocytes by immunofluorescence staining or electron microscopy, in 13 of 15 patients studied. High dimensional analyses of GI tissues revealed low levels of inflammation in general, including active downregulation of key inflammatory genes such as *IFNG, CXCL8, CXCL2* and *IL1B* and reduced frequencies of proinflammatory dendritic cell subsets. To evaluate the clinical significance of these findings, examination of two large, independent cohorts of hospitalized patients in the United States and Europe revealed a significant reduction in disease severity and mortality that was independent of gender, age, and examined co-morbid illnesses. The observed mortality reduction in COVID-19 patients with GI symptoms was associated with reduced levels of key inflammatory proteins including IL-6, CXCL8, IL-17A and CCL28 in circulation but was not associated with significant differences in nasopharyngeal viral loads. These data draw attention to organ-level heterogeneity in disease pathogenesis and highlight the role of the GI tract in attenuating SARS-CoV-2-associated inflammation with related mortality benefit.

**One Sentence Summary:** Intestinal infection with SARS-CoV-2 is associated with a mild inflammatory response and improved clinical outcomes.

## Introduction

Coronavirus disease 2019 (COVID-19), is a multisystem illness caused by severe acute respiratory syndrome coronavirus 2 (SARS-CoV-2) a recently discovered novel betacoronavirus(*1-3*). Manifestations of COVID-19 range from asymptomatic infection to severe, life-threatening disease with end-organ damage(*3-5*). Common symptoms of COVID-19 include fever, cough, and shortness of breath that occur within 2 to 14 days after exposure to SARS-CoV-2(*6*). A subset of COVID-19 patients also report gastrointestinal (GI) symptoms, comprising nausea, vomiting, diarrhea, abdominal pain and/or loss of appetite(*7-11*), which is not surprising given that the host receptor for SARS-CoV-2 (ACE2 receptor) is highly expressed on intestinal epithelium(*12, 13*). Several studies have demonstrated the presence of SARS-CoV-2 RNA in the fecal samples of infected persons(*11, 14-17*) and in some cases even after clearance from respiratory samples(*17*). Additionally, several members of the *Coronaviridae* family are known to be enterotropic, causing gastroenteritis in addition to respiratory illness(*18*). Notably, SARS-CoV and MERS-CoV, closely related to SARS-CoV-2, have been identified in fecal samples of infected individuals(*19-22*). Finally, the presence of GI involvement by SARS-CoV-2 has also been suggested by epidemiological(*23*), clinical(*24*), non-human primate(*25*) and *in vitro*(*26-29*) data. However, to date, there is limited evidence of SARS-CoV-2 infection of human enterocytes(*15*) and there are no studies on the responses of the GI immune system, arguably the largest in the body, in COVID-19 patients.

Immune dysregulation has been suggested as a primary driver of morbidity and mortality in COVID-19. Several cytokines and other immunological parameters have been correlated with COVID-19 severity. Most notably, elevated IL-6, IL-8, IL-10, MCP-1 and IP-10 levels were detected in hospitalized patients, especially critically ill patients, in several studies, and were associated with ICU admission, respiratory failure, and poor prognosis(*3, 30-36*). Alongside this pro-inflammatory cytokine environment, significant immune cell alterations have been described such as lymphopenia, T and B cell activation, and exhaustion(*37*), as well as altered frequencies of myeloid cells, including conventional dendritic cells (cDCs) and plasmacytoid dendritic cells (pDCs)(*5, 38, 39*).

Given the emerging evidence of enteric involvement by SARS-CoV-2, immune dysregulation in COVID-19 and the propensity of the GI immune system to suppress inflammation, we aimed to document infection of the GI tract in patients with COVID-19 and to define the impact of GI involvement on disease pathogenesis. Here, we present findings from well-characterized cohorts of COVID-19 patients hospitalized in tertiary care centers, from both New York City, USA and Milan, Italy, where we conducted high dimensional analyses of mucosal and systemic immune parameters and investigated disease outcomes associated with GI involvement in COVID-19 patients.

## Results

### The gastrointestinal tract of SARS-CoV-2 infected patients was endoscopically uninflamed in most patients

We sought to obtain GI tissue to determine if SARS-CoV-2 virus can be detected in human enterocytes *in vivo* as suggested by *ex vivo* enteroid studies(*40, 41*). To this end, we enrolled 18 COVID-19 patients and 10 SARS-CoV-2 uninfected controls who underwent upper GI endoscopy (SARS-CoV-2 infected n=16, uninfected n=8), colonoscopy (SARS-CoV-2 infected n=1, uninfected n=1) or both upper endoscopy and colonoscopy (SARS-CoV-2 infected n=1, uninfected n=1) (Table 1 and table S1). Patient 10 was initially suspected to have COVID-19 but was ultimately excluded after multiple negative SARS-CoV-2 nasopharyngeal (NP) PCR tests and negative COVID-19 antibody tests. The remaining 17 cases were classified as asymptomatic / mild / moderate (n=10) or severe (n=7) disease according to the criteria detailed in table S2. GI biopsies were performed after 17.3 ± 17.5 days from last positive nasopharyngeal (NP) swab (if patient had a positive NP swab after the procedure, it was considered to be 0 days from last PCR positive) (Fig. 1A). COVID-19 symptoms on presentation and treatment regimens were diverse as detailed in table S3. Sample allocation for different assays is detailed in fig. S1 and table S4.

**Table 1.** Clinical characteristics of patients who underwent endoscopic GI biopsies.

| Patient # | Age* | Sex | Procedure | Tissue sample location | COVID-19 Antibody | Days from symptom onset (or first SARS-CoV-2 PCR if asymptomatic) to procedure | Days from last positive SARS-CoV-2 PCR to procedure (0 if positive swab after procedure) | COVID severity on admission |
| --- | --- | --- | --- | --- | --- | --- | --- | --- |
| 1 | 60-65 | M | EGD | Duodenum | NA | 10 | 10 | Severe with EOD |
| 2 | 5-10 | M | EGD & colonoscopy | Intestinal Transplant | Positive | 23 | 0 | Severe |
| 3 | 75-80 | M | EGD | Duodenum | NA | 25 | 9 | Mild |
| 4 | 30-35 | F | EGD | Duodenum | NA | 44 | 31 | Severe with EOD |
| 5 | 60-65 | M | EGD | Duodenum | Positive | 24 | NA | Moderate |
| 6 | 50-55 | M | EGD | Duodenum | Positive | 11 | 0 | Asymptomatic / mild |
| 7 | 55-60 | M | EGD | Duodenum | Positive | 19 | 19 | Asymptomatic / mild |
| 8 | 65-70 | F | EGD | Duodenum | Positive | 30 | 23 | Severe |
| 9 | 75-80 | M | EGD | Duodenum | Positive | 24 | 0 | Severe |
| 10** | 80-85 | F | EGD | Duodenum | NA | NA | NA | NA |
| 11 | 90-95 | F | EGD | Duodenum | Positive | 36 | 0 | Severe with EOD |
| 12 | 50-55 | F | Colonoscopy | Ileum | NA | 37 | 31 | Mild |
| 13 | 40-45 | M | EGD | Duodenum | Positive | 18 | 0 | Mild |
| 14 | 65-70 | F | EGD | Duodenum | Positive | 37 | 32 | Severe with EOD |
| 15 | 30-35 | F | EGD | Duodenum | Positive | 50 | 48 | Mild |
| 16 | 70-75 | F | EGD | Duodenum | Positive | 25 | 19 | Mild |
| 17 | 50-55 | M | EGD | Duodenum | NA | 52 | 52 | Mild |
| 18 | 35-40 | M | EGD | Duodenum | NA | 3 | 3 | Asymptomatic / mild |

\* Age is provided as a range to obscure identifying information related to individuals
\*\*Patient 10 was excluded from analysis because SARS-CoV-2 nasopharyngeal PCR testing was repeatedly negative and her COVID-19 antibodies were also negative. Her inclusion initially was based on an atypical pulmonary infiltrate on chest X-ray and a viral syndrome in two of her family members.
Abbreviations: EGD = Esophagogastroduodenoscopy, F = Female, M = Male, EOD = End Organ Damage, NA = Not Available

**Fig 1.**
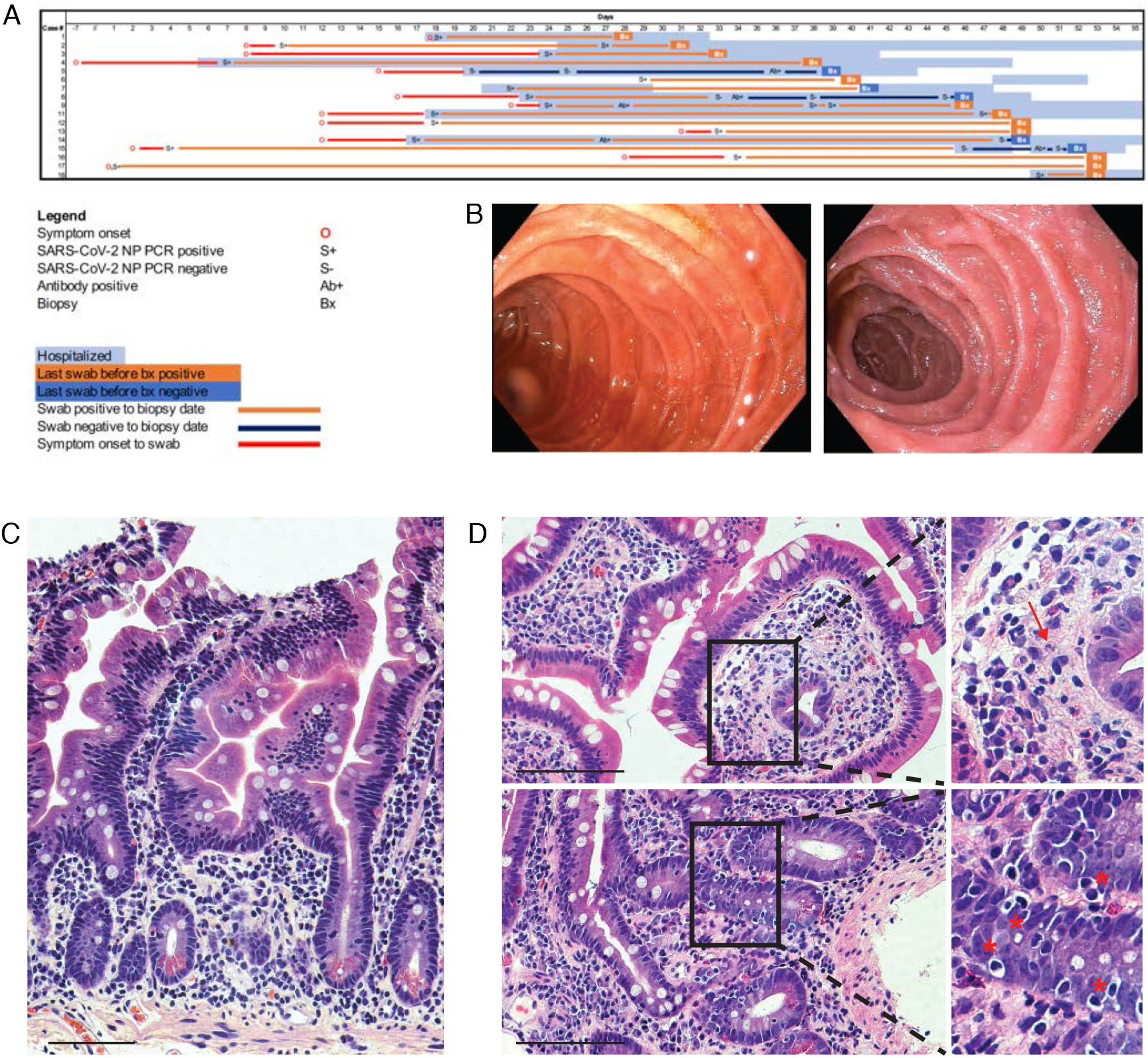
Clinical timing, endoscopic findings and histologic features in the small intestines of COVID-19 patients. (**A**) Timing of GI evaluation with respect to COVID-19 disease course. (**B**) Representative endoscopic images of the duodenum in COVID-19 (left) and control (right) patients. (**C**) Histologically normal duodenal tissue in a COVID-19 patient. (**D**) Histologic signs of inflammation detected in duodenal biopsies of COVID-19 patients including neutrophils (arrow) and increased intraepithelial lymphocytes (*). Scale bar; 100 µm.

The GI mucosa was endoscopically uninflamed in all subjects regardless of the severity of illness, with no evidence of loss of vascularity, edema, friability, erosions or ulcerations (Fig. 1B), except for one post-intestinal transplant case where inflammation was attributed to transplant rejection. Histopathological examination revealed normal histology in 4 cases, while of the remaining cases, we observed a mild increase in intraepithelial lymphocytes (IELs) in 8 cases and a scant neutrophilic infiltrate in 7 cases (Fig. 1, C and D, fig. S2 and, table S4).

### Small bowel enterocytes have robust expression of Angiotensin converting enzyme-2 (ACE2) and harbor SARS-CoV-2 antigens

An initial step in SARS-CoV-2 pathogenesis entails binding of viral spike (S) protein to the host ACE2 receptor, leading to viral entry and infection(*42*). Using immunofluorescence (IF) staining, we observed robust and extensive expression of ACE2 on the small intestinal brush border in both controls and COVID-19 patients (Fig. 2, A to H). Additionally, we detected SARS-CoV-2 nucleocapsid protein in small intestinal enterocytes of COVID-19 patients (Fig. 2, I and N, fig. S3), but not controls (Fig. 2, M and R, fig. S4), indicative of virus infection in these cells. Remarkably, when present, the distribution of viral antigens was patchy in the upper small intestines (duodenum; Fig. 2, I to L), but diffuse in the lower small intestines (ileum; Fig. 2, N to Q). Positive staining was exclusively seen in the epithelium irrespective of intestinal location of the biopsies. Overall, of the 11 COVID-19 patients where IF staining was performed, 10 showed viral antigen on IF microscopy in at least one intestinal segment (duodenum or ileum) (table S4). Interestingly, the presence of viral antigens on IF did not correlate with the presence of histologic abnormalities. As negative controls, 5 duodenal biopsies and 6 ileal biopsies from 10 patients collected prior to the pandemic (table S5), showed no evidence of viral antigens on immunostaining (Fig. 2, M and R, fig. S4).

**Fig 2.**
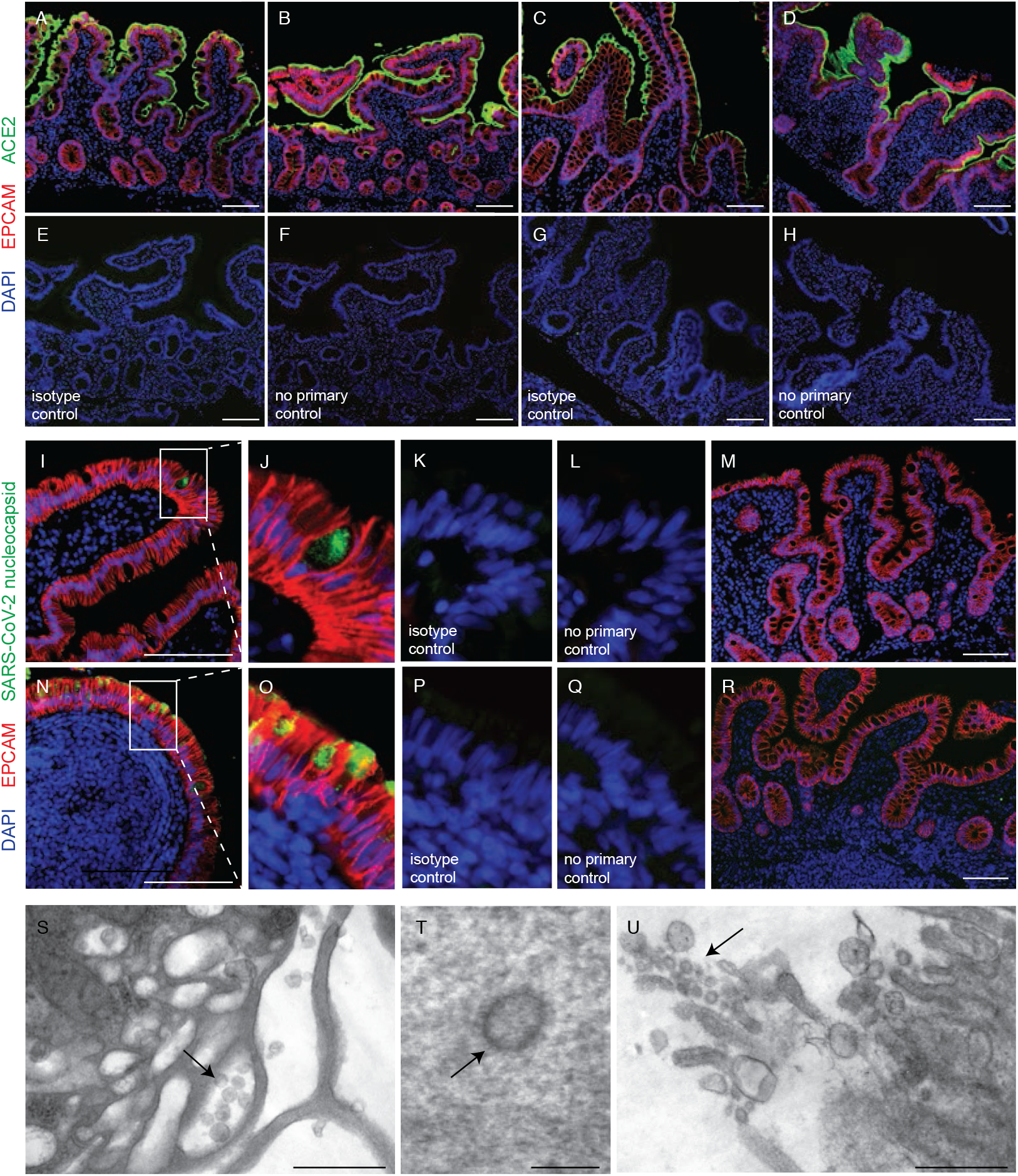
SARS-CoV-2 viral particles and protein are detectable in intestinal tissues of COVID-19 patients. (**A-H**) Immunofluorescent staining of duodenal (**A, B**) and ileal (**C, D**) biopsies of COVID-19 patients (**B, D**) and controls (**A, C**) with ACE2 (green), EPCAM (red) and DAPI (blue) including isotype (**E, G**) and no primary (**F, H**) controls. (**I-R**) Immunofluorescent staining of duodenal (**I-M**) and ileal (**N-Q**) biopsies from COVID-19 patients (**I-L, N-Q**) and controls (**M, R**) with SARS-CoV-2 nucleocapsid (green), EPCAM (red) and DAPI (blue) including isotype (**K, P**) and no primary (**J, Q**) controls. (**S-U**) Electron microscopy of duodenal biopsies (**S, T**) and an ileal biopsy (**U**) from COVID-19 patients showing viral particles (arrows). Scale bars; 100 µm (**A-R**), 0.5 µm (**S**,**U**), 0.1 µm (**T**).

### Ultrastructural analyses of GI tissues reveal viral particles in small intestinal enterocytes

Transmission electron microscopy of intestinal biopsy tissues revealed the presence of 70-110 nm viral particles in the enterocytes of the duodenum and ileum (Fig. 2, S to U). Pleomorphic, spherical structures, morphologically consistent with SARS-CoV-2 viral particles were observed in conjunction with small vesicles within enterocytes along the basolateral surface of the enterocytes (Fig. 2S) and blebbing off of the enterocyte apex (Fig. 2U). Particles with distinct, stalk-like projections (corona) were seen within the enterocyte cytoplasm (Fig. 2T). Overall, of the 13 patient samples processed for electron microscopy studies, 6 showed viral-like particles on ultrastructural analyses (table S4).

### Infectious virions could not be isolated from the GI tissues of COVID-19 patients

Attempts were made to assess for the presence of potentially infectious virions in the intestines of COVID-19 patients. When we inoculated Vero E6 cells with the supernatants of homogenized intestinal tissues from COVID-19 patients, we did not observe any apparent cytopathic effects (CPE) in the cells, despite culturing them for a week. In addition, cell culture supernatants did not reveal the presence of viral RNA by quantitative RT-PCR (qRT-PCR) and plaque assays. Experiments showed no plaque formation in Vero E6 cells after staining with 2% crystal violet solution.

### GI lamina propria pro-inflammatory dendritic cells are depleted in COVID-19 patients

Next, we performed mass cytometry (CyTOF) based immunophenotypic analyses on the GI tissues of 13 and peripheral blood of 10 COVID-19 cases and 10 controls (Table 1, table S1, fig. S1). GI tissues were processed to separate lamina propria (LP) and epithelial compartment (EC) fractions and both compartments were analyzed separately. Immune populations were clustered on the basis of cell-type specific markers for both the intestinal compartments (LP and EC) and blood (Fig. 3, A, C and G, fig. S6A and S7A, data file S1). While the overall distribution of canonical immune cell subsets in the GI LP were comparable between COVID-19 and control patients (Fig. 3, A and B), few immune populations showed differences as detailed below. Additionally, no clear differences in the LP could be discerned between patients with asymptomatic/mild/moderate disease and those with severe disease (Fig. 3B, data file S2).

**Fig 3.**
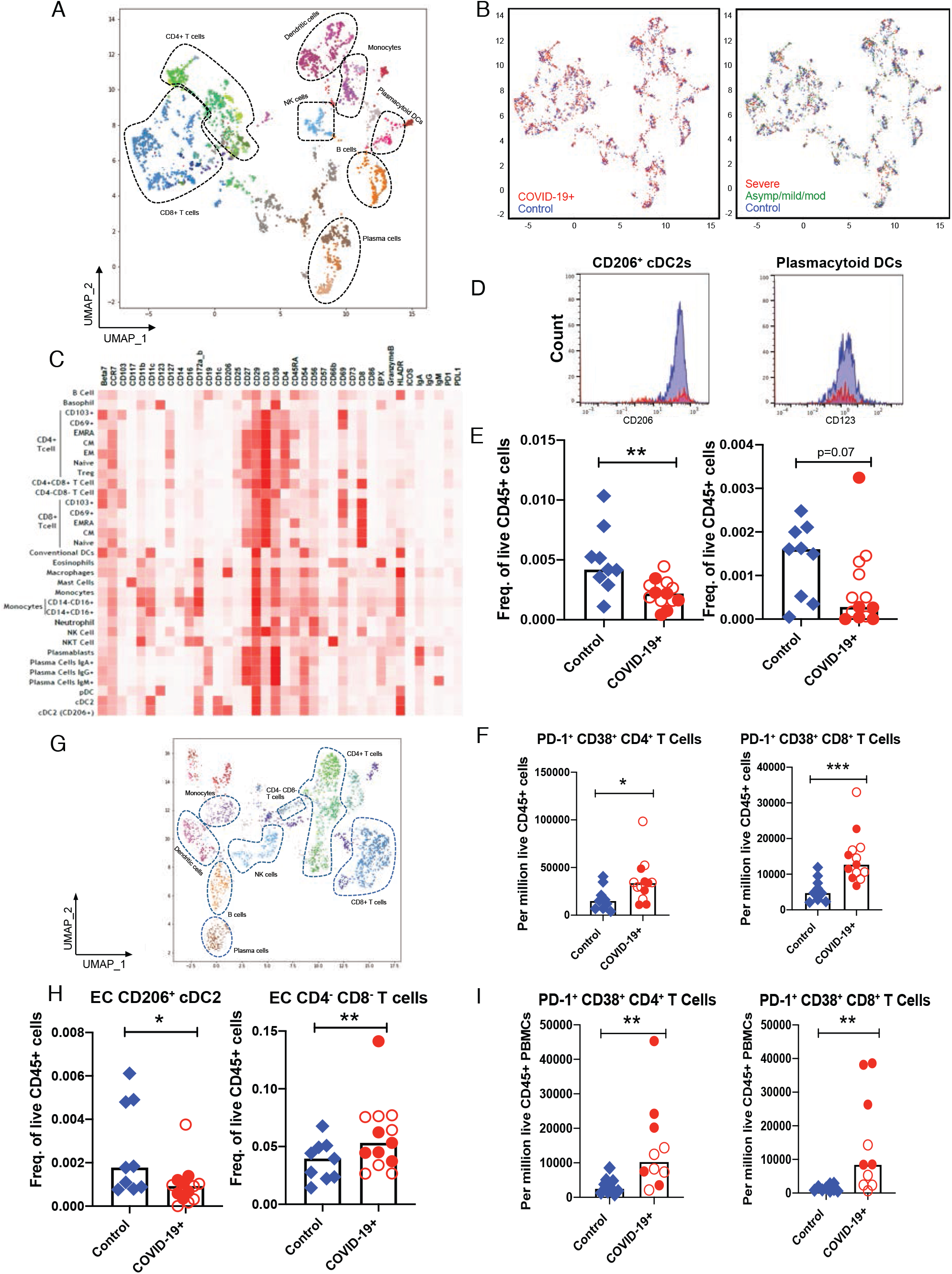
CyTOF-based analysis identified immune cell signatures in intestinal biopsies and blood from COVID-19 patients and controls. Uniform Manifold Approximation and Projection (UMAP) presentation of the eight clusters of lamina propria immune populations based on 38 markers (**A**), by infection status (**B**, left panel) with COVID-19 patients (red) and controls (blue), and by disease severity (**B**, right panel) with controls (blue), severe COVID-19 patients (red) and asymptomatic/mild/moderate COVID-19 patients (green). (**C**) The heatmap shows different lamina propria immune populations of COVID-19 patients and controls based on specific cell type markers. (**D**) The histograms show the decrease in the expression of CD206 and CD123 in dendritic cell populations of COVID-19 patients compared to the controls. (**E**) Relative frequencies of CD206+ cDC2 and plasmacytoid DCs in lamina propria of controls and COVID-19 patients (unsupervised analysis) (**F**) Relative frequencies of PD-1+ CD38+ (effector) CD4+ and CD8+ T cells in lamina propria of control and COVID-19 patients (supervised analysis) (**G**) UMAP presentation of the eight clusters of immune populations based on 38 markers in the epithelial compartment of intestinal biopsies. (**H**) Relative frequencies of CD206+ cDC2 subset of dendritic cells and CD4-CD8-T cells in the intraepithelial compartment of controls and COVID-19 patients (unsupervised analysis). (**I**) Relative frequencies of PD-1+ CD38+ (effector) CD4+ and CD8+ T cells in blood of controls and COVID-19 patients (supervised analysis). Open red circles denote patients with asymptomatic/mild/moderate disease while filled red circles denote patients with severe COVID-19. All bar plots represent median values. * p<0.05, **p<0.01, ***p<0.001.

In the LP, among myeloid cells, CD206^+^CD1c^+^ “inflammatory” cDC2 (conventional DCs)(*43*) were reduced in COVID-19 cases compared to controls (0.4-fold decrease, p=0.01). Additionally, plasmacytoid DCs (pDCs) were reduced in COVID-19 cases (0.5 fold decrease, p=0.07) (Fig. 3, D and E), analogous to changes described in the peripheral blood of COVID-19 patients(*36*). Among other LP populations, effector (PD-1^+^CD38^+^) CD4^+^ and CD8^+^ T cells were significantly increased in COVID-19 cases compared to the controls (Fig. 3F), while CD8^+^CD103^+^ T cells (tissue resident memory subset) were higher in COVID-19 cases compared to controls (1.7-fold increase, p=0.06) (fig. S5A). Neutrophils (3-fold), eosinophils (1.9-fold), CD4^-^CD8^-^T cells (3.5-fold) and CD4^+^CD103^+^ T cells (1.8-fold) were increased in COVID-19 cases, but these differences were not statistically significant possibly due to the sample size. We also observed a trend of non-significantly decreased regulatory CD4^+^ T (T_REG_) cells (0.6-fold decrease) and increased IgM^+^ plasma cells (2.3-fold increase) in the LP of infected cases vs controls (fig. S5A). The distribution of naïve and memory CD4^+^ and CD8^+^ T cells was altered in COVID-19 patients, with a reduction of naïve CD4^+^ T cells and EMRA (effector memory re-expressing RA) CD8^+^ T cells in the LP of COVID-19 patients (fig. S5B), but this difference did not reach statistical significance.

Similar to the LP, the EC showed a reduction of CD206^+^ cDC2 in COVID-19 cases compared to controls (0.4-fold decrease, p=0.05), while the CD4^-^CD8^-^ subset of IELs was significantly increased (1.6-fold-increase, p=0.03) (Fig. 3H). CD8^+^ T cells, the dominant IEL population, showed an increase (2.6-fold) in COVID-19 cases compared to controls but the difference did not reach statistical significance (p=0.4) (data file S2), likely owing to inter-patient variability, also observed by light microscopy. A subset of CD8^+^ IELs, CD8^+^CD69^+^ T cells, showed a non-significant increase in COVID-19 cases vs controls (3.9-fold, p=0.2), while plasma cells were comparable between the cases and controls (fig. S6, B and C).

In the peripheral blood, cell type assignments were carried out using specific cell surface markers (fig. S7A). We observed that effector (PD-1^+^CD38^+^) T cells (for both CD4^+^ and CD8^+^ T lymphocytes) were significantly increased in PBMCs of SARS-CoV-2 infected individuals (Fig. 3I). CD14^-^CD16^+^ inflammatory monocytes trended lower in COVID-19 cases compared to controls (0.4-fold decrease, p=0.09). In contrast, CD14^+^CD16^-^ “classical” monocytes were comparable in COVID-19 cases and controls (p=0.3) (fig. S7B). Additionally, a non-significant increase in IgG^+^ plasma cells (7.2-fold, p=0.13) and a non-significant decrease in T_REG_ (0.8-fold, p=0.2) were observed in COVID-19 cases (fig. S7, C and D, data file S2). Finally, a significant increase in activated (CD29^+^CD38^+^) CD4+ T cells was noted in the peripheral blood of COVID-19 cases compared to controls (fig. S8A) and a non-significant increase of these activated T cells in the LP of COVID-19 patients (fig. S8B). Details of all immune population changes in the intestinal tissues and in circulation are provided in data file S2.

Altogether, similar to published data from the peripheral blood of COVID-19 patients(*36*), intestinal tissues from COVID-19 cases showed reduced pro-inflammatory DCs and pDCs but increased effector T cells compared to controls.

### GI lamina propria pro-inflammatory pathways are downregulated in COVID-19 patients

To further probe the molecular response of the GI tract following SARS-CoV-2 infection, we performed RNA-Seq on the EC and LP intestinal compartments separately in 13 COVID-19 patients and 8 controls. Samples derived from the EC and LP clustered separately on the basis of their top transcriptional signatures, demonstrating distinctness of the two compartments within the GI tract (fig. S9, data file S3). Accordingly, comparisons between COVID-19 cases and controls were performed separately for each tissue, and 1063 differentially expressed genes (DEG) were identified out of total 11419 genes detected (Fig. 4A, data file S3). The majority of DEGs were detected in the LP (1061, false discovery rate (*42*) *≤* 0.05), compared to 12 DEGs in the EC that largely overlapped with the LP (Fig. 4A). Both LP and EC showed upregulation of genes involved in immunomodulation, including the anti-microbial peptide *LCN2*, and the metallothioneins *MT1E, MT1F, MT1H, MT1M, MT1X, MT2A* and *TMEM107*. In addition, heat shock proteins, *HSPA1A* and *HASPA1B*, were downregulated in both tissues. Pathway enrichment analysis of DEGs ranked by significance revealed several KEGG pathways that were depleted in COVID-19 patients compared to controls (Fig. 4B). Downregulation of pathways linked to T_H_17 cell differentiation and inflammatory bowel diseases (IBD) was characterized by the depletion of *RORA, IL4R, IFNG, IL18R1, IL1B, STAT4* and *HLA-DRA*. Pathways linked to antigen processing, T_H_1 and T_H_2 cell differentiation, and MAPK signaling were significantly downregulated in the LP from COVID-19 patients. In contrast, genes associated with metabolic functions, including amino acid metabolism (*NOS2, SMS, ALDH2, GOT2*), mineral absorption (*MT1G, MT2A, MT1E*), as well as mucin biosynthesis (*GALNT7, GALNT3, GALNT8*) were significantly upregulated in COVID-19 patients compared to controls (Fig. 4B).

**Fig 4.**
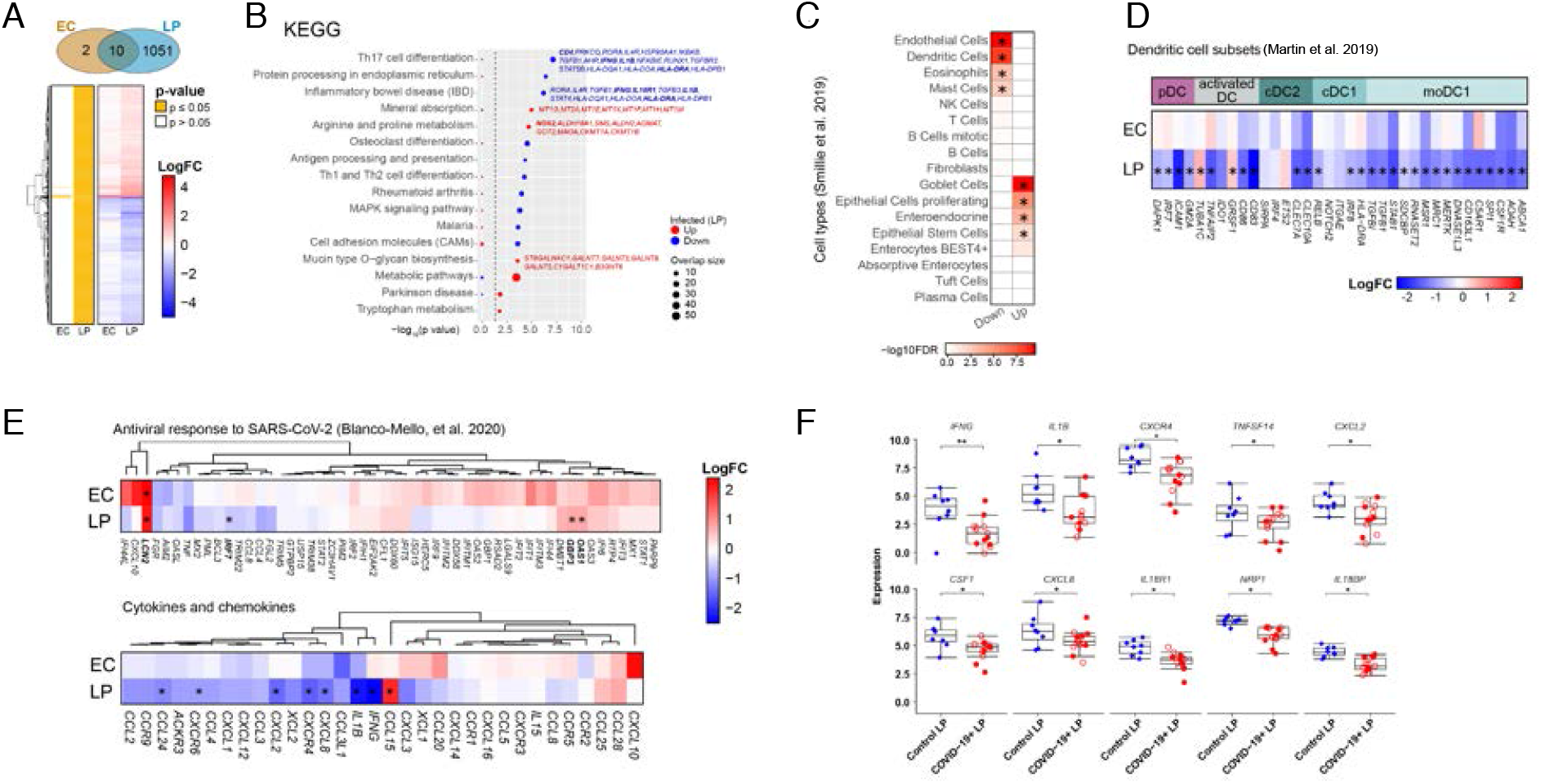
Transcriptional changes in intestinal biopsies from COVID-19 patients compared with controls. (**A**) Hierarchical clustering of average expression changes for 1,063 genes (rows) with induced (red) or depleted (blue) expression (FDR ≤ 0.05) in the epithelial compartment (EC) and lamina propria (LP) fractions of intestinal biopsies from COVID-19 patients. The panel on the left indicates significant genes for each tissue fraction in yellow. The color bar indicates the average log2 fold-change (FC). (**B**) Top enriched pathways (KEGG) that are induced (red) or depleted (blue) in LP of COVID-19 patients are displayed. The dash line indicates the p ≤ 0.05 cutoff. Gene names are indicated for main pathways related to inflammation and cell type composition. The dashed line indicates the p ≤ 0.05 cutoff. (**C**) Deconvolution of main gastrointestinal cell types enriched or depleted in the LP of COVID-19 patients against controls. Reference scRNA-seq cell type signatures were taken from *Smillie et al. 2019*. (p ≤ 0.05, Fisher’s exact test). (**D**) Average expression changes for dendritic cell markers in the LP and EC compartments. Reference scRNA-seq cell type signatures were taken from *Martin et al. 2019*. The color bar indicates the average log2 fold-change (FC). (**E**) Hierarchical clustering of average expression changes (columns) in the EC and LP fractions for genes related to antiviral response to SARS-CoV-2 in post-mortem lung tissue of COVID-19 patients as described by *Blanco-Mello et al*. 2020 (top panel) and for cytokines and chemokines (bottom panel). Significant genes are indicated by the asterisks. The color bar indicates the average log2 fold-change (FC). (**F**) The gene expression levels for the top 10 significant chemokines and cytokines in the LP of COVID-19 patients (n=13) and controls (n=8). *p < 0.05, **p < 0.01.

We considered the possibility that the observed expression changes could imply alterations in relative cell type proportions (in addition to transcriptional alterations within cells). Therefore, we interrogated GI data derived from single-cell RNA-seq(*44*) for enrichment of cell type-specific gene expression signatures. Consistent with our CyTOF data (Fig. 3 and data files S1 and S2), genes associated with DCs and eosinophils were reduced in COVID-19 patients compared to controls (Fig. 4C). Additionally, signatures related to the size of endothelial cell and mast cell pools were reduced, while genes linked to goblet cells, proliferating epithelial cells, enteroendocrine cells and epithelial stem cells were increased, possibly reflecting the sequelae of intestinal epithelial infection by SARS-CoV-2 and subsequent recovery (Fig. 4C).

Given the unexpected reduction in DC numbers in the GI tissues, we probed myeloid gene signatures further, and found significant downregulation of genes associated with pDC *(DAPK1, IRF7, ICAM1* and *GM2A*), activated DCs (*TNFAIP2, CD86, CD83*), cDC1 (*RELB, IRF8* and *HLA-DRA*) and cDC2 (*CLEC7A* and *CLEC10A*). Of note, we also found that LP genes associated with inflammatory DCs (monocyte-derived DCs, MoDCs) (*TGFBI, TGFB1, STAB1, SDCBP, RNASET2, MSR1, MRC1, MERTK, DNASE1L3, CD163L1, C5AR1, SPI1, CSF1R, AOAH, ABCA*) were significantly reduced (Fig. 4D), which was consistent with the reduced number of inflammatory DCs observed in our CyTOF results.

Finally, we looked at the average EC and LP expression of recently reported gene signatures linked to the antiviral response against SARS-CoV-2 from post-mortem lung tissue samples(*30*), and human intestinal organoids(*29*). Although we did not observe a substantial acute SARS-CoV-2 response, there was significant upregulation of *LCN2* in both EC and LP, and *OAS* and *GBP3* in the LP only. Notably, we did observe a trend towards induction of antiviral response genes in the EC, where expression of canonical antiviral genes such as *IFI44L, IFIT1, IFITM3, IFI44, IFI6* and *OAS3* was increased (Fig. 4E, top panel).

Next, using gene set enrichment analysis (GSEA), we rank ordered the EC DEGs according to effect size (logFC * -logPvalue) and tested for enrichment in the reported SARS-CoV-2 infected organoid gene signatures(*29*) (fig. S10A). The genes we found upregulated in the EC of COVID-19 patients were significantly enriched in the SARS-COV-2 infected organoid gene datasets. We then carried out Hallmark pathway enrichment analyses on this ranked EC gene list and found that the top two processes associated with genes upregulated in EC were interferon alpha response (normalized enrichment score (NES) 1.91, FDR<0.005) and interferon gamma response (NES=1.8, FDR=0.005) (fig. S10B). This enrichment is indicative of the host antiviral response against SARS-CoV-2 in the intestinal EC of COVID-19 patients.

We projected our RNA-seq dataset on the published data set from the human bronchial epithelial cells infected with SARS-CoV-2 and evaluated cytokines and chemokines in intestinal samples(*30*). Remarkably, we found that many of the inflammatory cytokines and chemokines such as IL-1*β*, IFN-*γ*, CCL24 and CXCL8 were downregulated in the intestines of COVID-19 patients (Fig. 4E, bottom panel). The only chemokine significantly increased was CCL15 which is reported to be structurally similar to antimicrobial peptides and has a role in maintaining intestinal homeostasis(*45*) (Fig. 4E, bottom panel). We also noted that the key inflammatory genes including *IFNG, IL1B, CXCR4, TNFSF14, CXCL2, CSF-1, CXCL8, IL18R1, NRP1 and IL18BP* were downregulated in intestinal LP of COVID-19 cases compared to the uninfected controls (Fig. 4F). Interestingly, the expression of NRP1, a recently discovered host factor for SARS-CoV-2 infection(*46*), was found to be significantly reduced in LP of COVID-19 patients compared to the controls.

Together, these data reveal a dynamic remodeling of GI tissues by SARS-CoV-2, notably with a significant downregulation of pathways associated with inflammation and antigen presentation in the LP, while with a concomitant activation of viral response signaling genes in the EC.

### Examination of a cohort of COVID-19 patients (Discovery Cohort) to assess the clinical impact of GI involvement: clinical characteristics

Given that GI biopsies from COVID-19 patients demonstrated a paucity of cellular inflammation and in some instances, a down regulation of key inflammatory genes, despite the presence of SARS-CoV-2 in the enterocytes of the respective patients, we hypothesized that intestinal involvement in COVID-19 would be associated with a milder disease course. To test our hypothesis, we examined a large, retrospective cohort of patients (henceforth referred to as the ‘Discovery Cohort’) who were hospitalized at Mount Sinai Hospital (MSH) between April 1, 2020 and April 15, 2020. Among 925 patients hospitalized with the diagnosis of COVID-19 during this period, after an initial screening, 634 cases met our inclusion criteria of age > 18 years and having a multiplexed cytokine panel performed during their admission (fig. S11). Demographics (gender, age and race/ethnicity) and clinical variables including the presence of comorbid diagnoses, COVID-19 severity and GI symptoms were carefully analyzed and documented. The mean age of patients was 64 ± 16 years (range 23-99 years); 28% of patients self-reported as Hispanic, 25% as African American, 22% as White and the remainder as Asian or other. Patients had multiple comorbid illness that included obesity (37%), hypertension (HTN) (36%) and diabetes mellitus (DM) (22%) (table S6). With regards to comorbid illnesses, the composition of the Discovery Cohort is similar to prior reports(*47*).

Next, we recorded the presence of GI symptoms that have been associated with COVID-19(*7-11*) in each of the patients. We only considered the GI symptoms that were present at the time of hospital admission (to avoid iatrogenic confounders) and focused on symptoms that were indicative of direct gastrointestinal involvement such as diarrhea, nausea and vomiting. We did not consider symptoms like anorexia and weight loss (which might be multifactorial), or dysgeusia and anosmia (indicative of nasopharyngeal involvement). Using these criteria, 299 patients (47%) reported any of the GI symptoms of interest (nausea, vomiting and/or diarrhea) with diarrhea being the most common (245 patients, 39%), followed by nausea (157 patients, 25%), and then vomiting (82 patients, 13%) (table S6).

### COVID-19 severity was significantly reduced in patients with GI symptoms when compared to those without GI symptoms on multivariate testing, accounting for age and comorbidities

At the outset, we performed univariate analyses comparing the distribution of COVID-19 severities in patients with and without GI symptoms. Overall, 54 (9%) patients had mild disease, 361 (57%) moderate, 158 (25%) severe and 61 (10%) had severe COVID-19 with end organ damage (EOD) (disease severity defined in Materials and Methods and table S2; table S6). During hospitalization, 110 patients were admitted to the ICU (17%) and 151 patients (24%) died by the end of data collection (6/15/2020) (table S6). Patients presenting with GI symptoms had less severe disease than patients without GI symptoms (p<0.001 Chi-square test, Table 2). Importantly, mortality was significantly lower in COVID-19 patients with GI symptoms (15.7%) than those without GI symptoms (31.0%; p<0.0001 Fisher’s exact test) (Table 2). Furthermore, each individual GI symptom (nausea, vomiting and diarrhea) was associated with less severe disease (p<0.02 Fisher’s exact test) and lower mortality (p<0.001 Fisher’s exact test) (table S7). These findings were further emphasized by Kaplan-Meier estimates of survival over short-term follow-up of 25 days (p<0.001 log-rank test) (Fig. 5A and fig. S12, A and B). Consistent with prior reports(*35, 47, 48*) older age and higher disease severity were associated with higher mortality in the Discovery Cohort (table S8), providing validity to our findings.

**Table 2.**
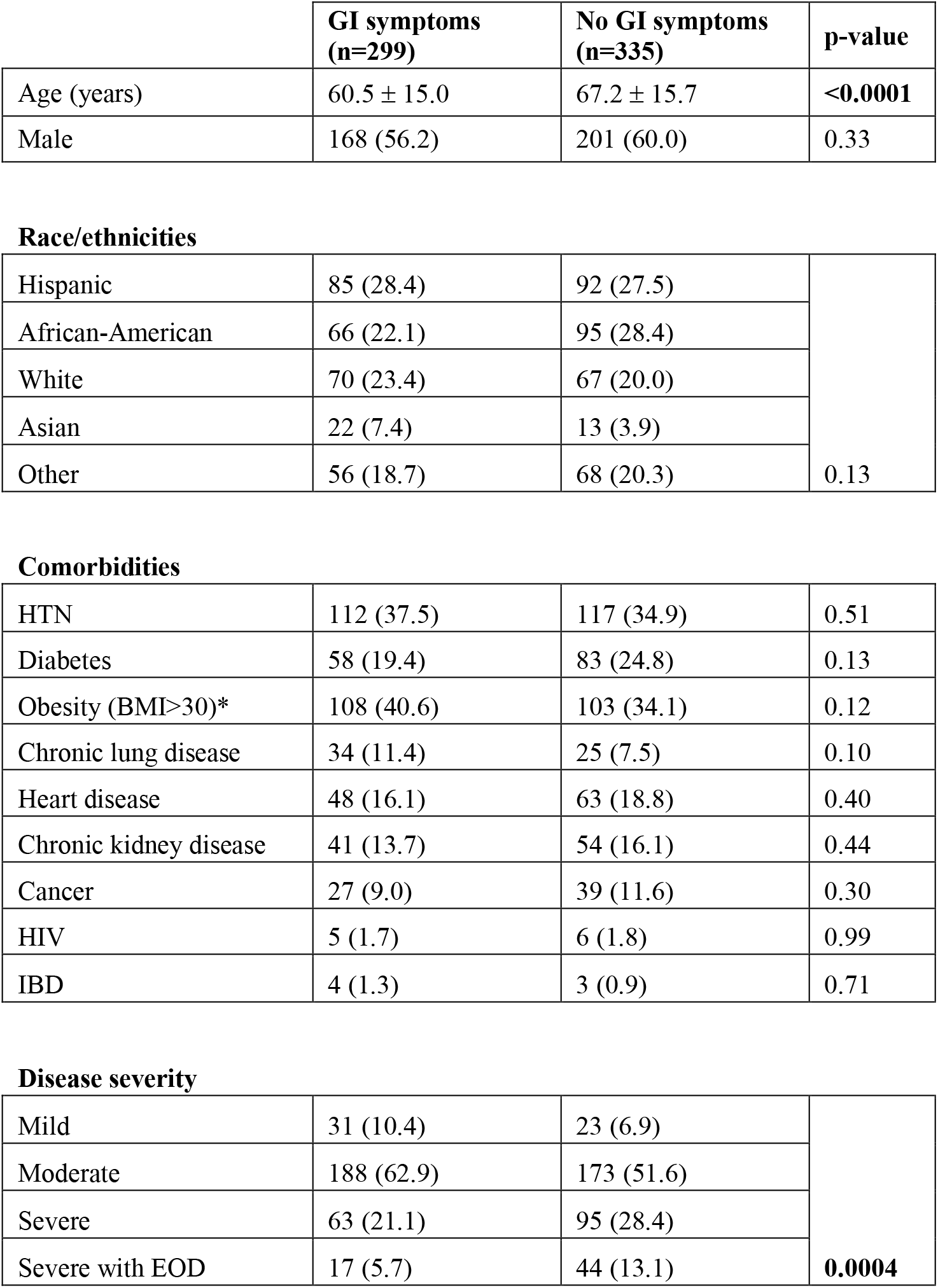
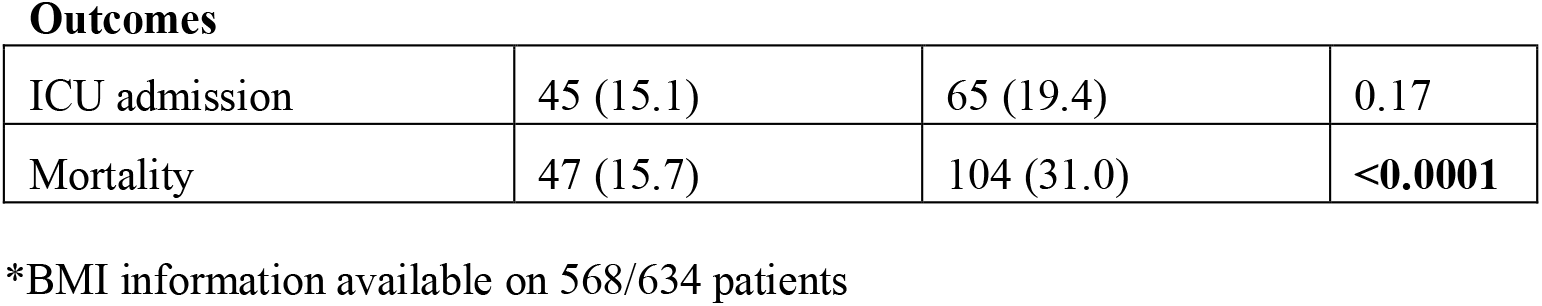
Basic demographics, clinical characteristics and outcomes in patients with and without GI symptoms. For age, the mean ± standard deviation is listed and an unpaired two-tailed t-test was performed. For categorical variables, the number of patients followed by the percent of patients in parentheses is listed and the Fisher’s exact test or the Chi-square test was used as appropriate.

|  | **GI symptoms**<br>**(n=299)** | **No GI symptoms**<br>**(n=335)** | **p-value** |
| --- | --- | --- | --- |
| Age (years) | 60.5 ± 15.0 | 67.2 ± 15.7 | **<0.0001** |
| Male | 168 (56.2) | 201 (60.0) | 0.33 |
| **Race/ethnicities** |  |  |  |
| Hispanic | 85 (28.4) | 92 (27.5) | 0.13 |
| African-American | 66 (22.1) | 95 (28.4) |  |
| White | 70 (23.4) | 67 (20.0) |  |
| Asian | 22 (7.4) | 13 (3.9) |  |
| Other | 56 (18.7) | 68 (20.3) |  |
| **Comorbidities** |  |  |  |
| HTN | 112 (37.5) | 117 (34.9) | 0.51 |
| Diabetes | 58 (19.4) | 83 (24.8) | 0.13 |
| Obesity (BMI>30)* | 108 (40.6) | 103 (34.1) | 0.12 |
| Chronic lung disease | 34 (11.4) | 25 (7.5) | 0.10 |
| Heart disease | 48 (16.1) | 63 (18.8) | 0.40 |
| Chronic kidney disease | 41 (13.7) | 54 (16.1) | 0.44 |
| Cancer | 27 (9.0) | 39 (11.6) | 0.30 |
| HIV | 5 (1.7) | 6 (1.8) | 0.99 |
| IBD | 4 (1.3) | 3 (0.9) | 0.71 |
| **Disease severity** |  |  |  |
| Mild | 31 (10.4) | 23 (6.9) | **0.0004** |
| Moderate | 188 (62.9) | 173 (51.6) |  |
| Severe | 63 (21.1) | 95 (28.4) |  |
| Severe with EOD | 17 (5.7) | 44 (13.1) |  |
| **Outcomes** |  |  |  |
| ICU admission | 45 (15.1) | 65 (19.4) | 0.17 |
| Mortality | 47 (15.7) | 104 (31.0) | **<0.0001** |

\*BMI information available on 568/634 patients

**Fig 5.**
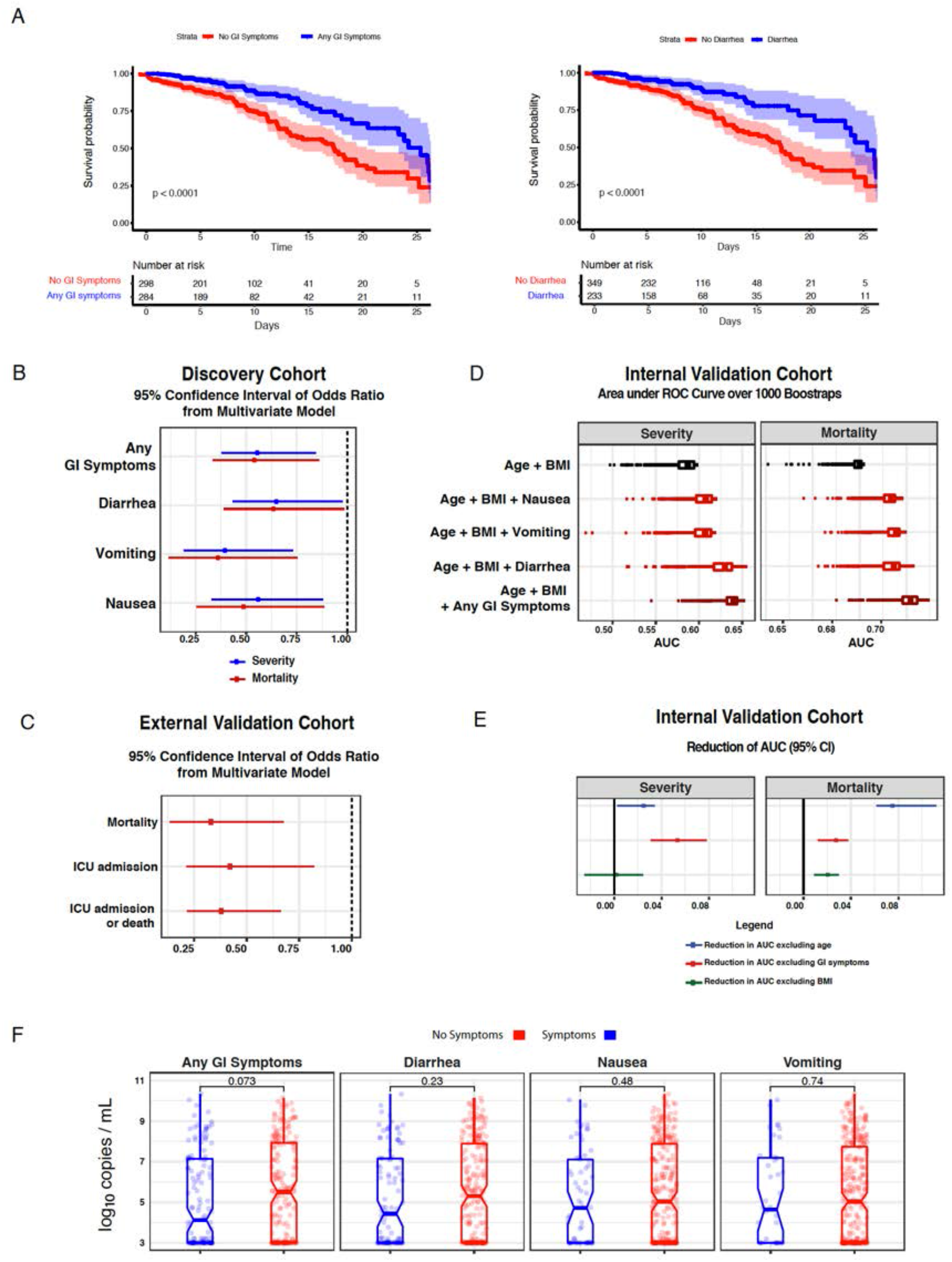
COVID-19 patients with GI symptoms had reduced severity and mortality despite similar nasopharyngeal viral loads compared to those without GI symptoms. (**A**) Kaplan-Meier (KM) curves for survival stratified by any GI Symptoms (left panel) and diarrhea (right panel) for patients in the Discovery Cohort. P-values from log-rank test and 95% confidence intervals of KM curves are shown. Below each KM curve, the number of patients at risk are reported for the respective time points. (**B**) Confidence intervals of odds ratio (95%) of GI symptoms based on 1000 bootstrap iterations in a multivariate logistic regression for severity (blue) and mortality (red). (**C**) Validation based on the External Cohort. Confidence intervals of odds ratio (95%) of diarrhea covariate based on 1000 bootstrap iterations to capture mortality, ICU admission and composite outcome of ICU admission or death. Results are based on multivariate models after accounting for confounders such as BMI, age, gender, lung disease, heart disease and hypertension. (**D**) Validation based on the Internal Cohort. Boxplot of AUC over 1000 bootstrap iterations to predict mortality and disease severity in the Validation Cohort. (**E**) Confidence intervals of the reduction in AUC (95%) based on 1000 bootstrap iterations for the model “Age + BMI + Any GI Symptoms” after removing age (blue), GI symptoms (red) and BMI (green). (**F**) SARS-CoV-2 viral load copies per mL (log_10_ transformed based on N2 primer with the addition of a constant) stratified by GI symptoms. The box plot represents median and interquartile range. P-values from two-tailed unpaired t-tests are reported.

Next, we decided to account for multiple comorbidities and demographics in determining the impact of GI symptoms on COVID-19 outcomes. While patients with GI symptoms (61 ± 15 years) were younger than those without GI symptoms (67 ± 16 years), the distribution of race, ethnicity and co-morbid illnesses including obesity, HTN, DM and inflammatory bowel diseases (IBD) were comparable between those with and without GI symptoms (Table 2). To account for potential confounders, especially the age difference, we created a multivariate model where we adjusted for the effect of age, body mass index (BMI), gender, race, diabetes, HTN, chronic lung disease and heart disease on outcomes. Consistent with published literature(*49-52*) (age and BMI were positively associated with COVID-19 severity and mortality (table S9. Using this multivariate model, we analyzed the impact of GI symptoms on disease outcomes. The presence of any GI symptoms taken together, as well as diarrhea, nausea, and vomiting taken individually, were inversely associated with COVID-19 severity and mortality (Fig. 5B, table S9). Patients who presented with GI symptoms had 50% reduced odds of having severe disease (odds ratio of 0.56) and death from COVID-19 (odds ratio of 0.54), compared to the patients who presented without GI symptoms (Fig. 5B, table S9).

### An External Validation Cohort further confirms decreased mortality in COVID-19 patients with GI symptoms on multivariate testing

Next, we identified an external cohort of 287 well-characterized patients, distinct from the MSH Discovery Cohort (henceforth referred to as ‘External Validation Cohort’) from Milan, Italy, to determine the impact of GI symptoms on COVID-19 associated outcomes. In this cohort, GI symptoms on admission were characterized as presence or absence of diarrhea (table S10). Consistent with the Discovery Cohort, patients with diarrhea on admission had significantly lower mortality (10.0%) when compared to patients without diarrhea (23.7%, p=0.008). Additionally, patients with diarrhea had lower composite outcome of mortality or ICU admission compared to those without diarrhea (20% vs 40%, p=0.001) (table S10). When we performed multivariate logistic regression adjusting for age, gender, BMI, diabetes, chronic heart and lung disease and other confounders in our External Validation Cohort, we observed that, the presence of diarrhea on admission was found to be significantly inversely associated with mortality with a median odds ratio of 0.33 over 1000 bootstrap iterations (Fig. 5C). Our observations from this External Validation Cohort were in alignment with those from our Discovery Cohort.

### Presence of GI symptoms can be used to predict reduced disease severity and mortality in patients with COVID-19

After observing significantly reduced mortality in COVID-19 patients with GI symptoms in our Discovery and External Validation Cohort, we developed a predictive model based on the Discovery Cohort and applied it to a distinct Internal Validation Cohort comprising 242 well-characterized patients with COVID-19, admitted between April 16, 2020 and April 30, 2020 to MSH. The inclusion of ‘any GI symptoms’ to a model consisting of age and BMI at baseline, improved the ability to predict severity and mortality with a median area under the curve (AUC) of 0.59 (age + BMI) vs. 0.64 (age + BMI + any GI symptoms) for disease severity and 0.70 (age + BMI) vs. 0.73 (age + BMI + any GI symptoms) for mortality (Fig. 5D, table S11). In addition, the effect of GI symptoms, age and BMI on the AUC was evaluated by excluding each variable one at a time from the model and calculating the consequent reduction in AUC. The exclusion of GI symptoms resulted in a significant reduction in AUC with a median value of 0.054 for disease severity and 0.03 for mortality. Notably, the effect of GI symptoms on the AUC was more dramatic than that of age (AUC reduction of 0.054 versus 0.025) for disease severity (Fig. 5E, table S11).

### Nasopharyngeal SARS-CoV-2 viral loads are similar in patients with and without GI symptoms

There is recent evidence that nasopharyngeal (NP) SARS-CoV-2 viral loads are correlated with disease outcomes(*53*), therefore, we compared NP viral loads in a subset of Discovery and Internal Validation Cohort patients (n=329, where these data were available). Patients with GI symptoms had similar SARS-CoV-2 NP viral loads compared to those without GI symptoms (mean log_10_ copies/mL 5.1 (SD 2.3) vs 5.6 (SD 2.4) respectively) (p=0.07); furthermore, no significant differences in viral loads were observed when analyzing each individual GI symptom (Fig. 5F). Therefore, the lower severity and mortality in those with GI symptoms are not explained by differences in viral loads.

### COVID-19 patients with GI symptoms have reduced levels of circulating cytokines associated with inflammation and tissue damage

To correlate the observed mortality difference between patients with and without GI symptoms with known biomarkers associated with severe COVID-19, we initially examined, a set of 4 cytokines, IL-6, IL-8, TNF-α, and IL-1β, measured on admission in all the patients as part of routine clinical care. IL-6, and IL-8 levels, known to be associated with poor survival(*35*) were found to be significantly reduced in circulation of patients with GI symptoms (FDR 10%) (fig. S13, table S12).

Next, to facilitate high dimensional analyses of potential immunological differences between patients with and without GI symptoms, we performed a validated, multiplexed proteomic assay (O-link), simultaneously quantifying 92 protein analytes in 238 patients (from among the Discovery and Internal Validation Cohorts; GI symptoms (n=104), no GI symptoms (n=134)) where serum samples were available for analyses. Unsupervised consensus clustering of these 92 analytes revealed six groups of analytes with similar expression patterns across all COVID-19 patients (Fig. 6A, table S13). Notably, analytes in clusters 5 and 6 displayed less correlation in patients with GI symptoms compared to those without GI symptoms (Fig. 6A, fig. S14). Next, we interrogated biological pathways over-represented in each cluster of soluble analytes. We found that the “KEGG Jak/Stat Signaling Pathway” was significantly enriched in Cluster 5; while the “Hallmark Inflammatory Response” pathway was significantly enriched in Cluster 4 (Fisher’s exact test 10% FDR). These pathways were downregulated in patients displaying diarrhea symptoms based on pathway level signatures (p<0.05 from t-test) (Fig. 6B); suggesting a reduced inflammatory response in patients affected by GI symptoms. In addition, we found that clusters 1, 2, 3, 5 and 6 were significantly downregulated in patients with GI symptoms compared to those without (FDR 15%) (Fig. 6C). This downregulation seemed to be driven mostly by diarrhea since the same clusters 1, 2, 3, 5 and 6 were significantly downregulated with FDR correction at 10% in patients who presented with diarrhea compared to those without symptoms. The reduced signal observed for nausea and vomiting might be due to reduced statistical power given by the smaller number of samples displaying vomiting (n=29) or nausea (n=54) symptoms.

**Fig 6.**
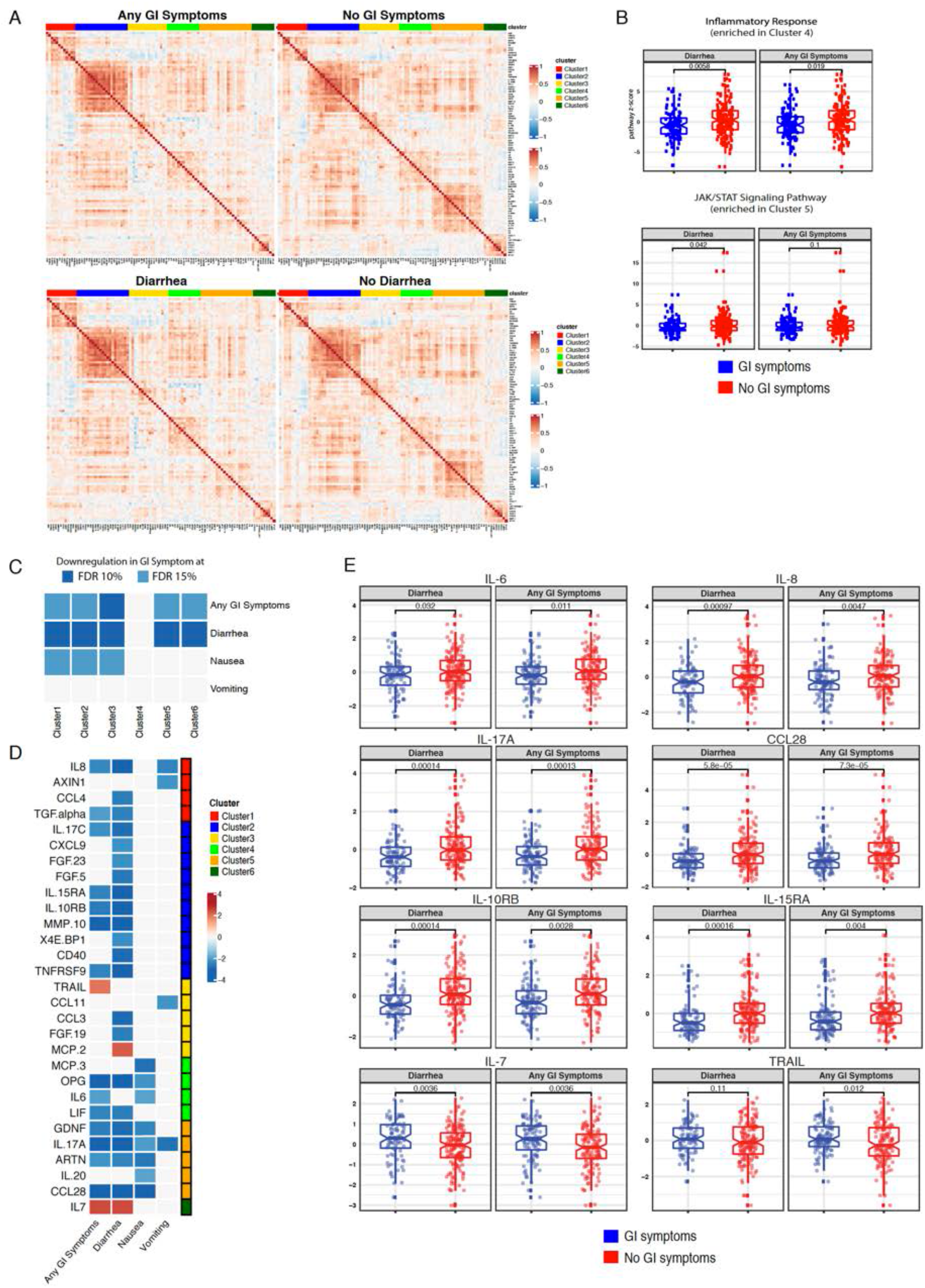
COVID-19 patients with GI symptoms have reduced levels of circulating inflammatory cytokines. (**A**) Correlation matrix (Pearson’s) for 92 markers contained in multiplexed proteomic platform (O-link) across patients with any GI symptoms (top left panel) compared with no GI symptoms (top right panel) and patients with diarrhea (bottom left panel) compared with patients without diarrhea (bottom right panel). Cluster assignment derived based on consensus clustering is reported on the top of the heatmap. (**B**) Boxplot of “Hallmark Inflammatory Response” and “KEGG JAK/STAT Signaling pathway” z-scores stratified by GI symptoms. P-values from unpaired t-test are reported. “Hallmark Immune Response” and “Hallmark JAK/STAT Signaling” pathways were found significantly enriched at 10% FDR in Cluster 4 and Cluster 5, respectively. (**C**) Association between proteomic clusters and GI symptoms, which was derived by comparing cluster signatures between asymptomatic and symptomatic groups via unpaired two-tailed t-test. Associations significant at 10% (dark blue) and 15% (light blue) FDR are reported. (**D**) Analytes associated with GI symptoms at 10% FDR based on unpaired t-test. The intensity of the color is proportional to the -log10 p-value. Negative associations are displayed in blue; while positive associations in red. On the right side of the heatmap, the cluster assignment for each marker is reported. (**E**) Boxplots represent median and interquartile range of select differentially expressed markers stratified by GI symptoms. P-values from unpaired t-test are reported.

When looking at each of the 92 analytes individually, key inflammatory cytokines and chemokines were significantly downregulated (IL-8, TGF-*α*, IL-17C, IL-15RA, IL-10RB, MMP10, TNFRSF9, OPG, IL-6, LIF, GDNF, IL-17A, ARTN and CCL28). On the other hand, TNF-Related Apoptosis Inducing Ligand (TRAIL), a cytokine with immune regulatory properties(*54, 55*) and IL-7, a cytokine associated with T cell development(*56*) were significantly upregulated in patients with GI symptoms compared to those without (t-test FDR 10%) (Fig. 6, D and E, table S14). When looking at each individual GI symptom, diarrhea had the most significantly differential analytes. Again, this difference between diarrhea, nausea and vomiting might be due to reduced statistical power given that fewer patients presented with nausea and vomiting than those who presented with diarrhea. Consistent with GI symptoms as a group, IL-7 was significantly increased, in addition, MCP-2 was significantly increased in patients presenting with diarrhea.

Thus, overall, GI symptoms are associated with significantly reduced levels of key inflammatory cytokines like IL-6, IL-8, IL-17 and CCL28 that are known to be associated with poor COVID-19 outcomes.

## Discussion

Given the robust expression of ACE2 on the small intestinal epithelium(*57*), at levels that are arguably among the highest in the body(*58*), we hypothesized that the GI tract would be susceptible to SARS-CoV-2 infection. This study was designed to test this hypothesis by examining for human intestinal infection by SARS-CoV-2. Herein, we report on the first detailed human study to demonstrate infection of intestinal enterocytes *in vivo* and to define the cellular and transcriptomic responses of GI tissue in COVID-19 patients. Having seen an unexpected mild inflammatory response in the intestinal tissues, despite evidence of SARS-CoV-2 proteins and viral particles, we next hypothesized that GI involvement could perhaps be associated with improved clinical outcomes. In two distinct and large cohorts of COVID-19 patients, we observed a significant reduction in mortality in patients with GI symptoms compared to those without GI symptoms, even after adjusting for multiple confounders including age and comorbid illnesses. These data identify a potentially novel aspect of COVID-19 pathogenesis and emphasize the presence of an ‘organ-specific’ program of host response to SARS-CoV-2, which in this case is associated with a survival advantage.

We considered several non-mutually exclusive hypotheses to explain the lower mortality in COVID-19 patients with GI symptoms. Multiple physiological mechanisms enable the intestines to maintain a state of “immunological tolerance”. Among these, IgA, the dominant intestinal immunoglobulin does not invoke the complement cascade unlike IgG, and effects a “non-inflammatory” pathogen neutralization. A recent study has demonstrated that dimeric Immunoglobulin A (IgA), that would be induced within mucosal tissues of the GI tract is significantly more potent than IgG or monomeric IgA in SARS-CoV-2 neutralization(*59*). Effective, but non-inflammatory viral neutralization at the intestinal mucosal surface would reduce tissue viral loads, potentially attenuating the inflammatory cascade. Additionally, we found increased expression of anti-inflammatory gene products, including transcripts involved in the biogenesis of the gut-specific MUC2 mucin, a mucus-forming glycoprotein released by goblet cells that contributes to intestinal tolerance(*60*). We also examined for differences in NP viral loads between patients with and without GI symptoms, hypothesizing that perhaps the virus in some patients is “diverted” away from the respiratory system. However, we did not observe significant NP viral load differences between patients with and without GI symptoms. Therefore, we believe that limited intestinal and systemic inflammation contributed to improved mortality in COVID-19 patients with GI involvement.

Multiple lines of evidence from our study and from the existing literature support this possibility. First, and most evident was the relative paucity of inflammatory infiltrates in the intestinal epithelium as well as LP of COVID-19 patients. Additionally, we observed a rather significant lack of cDC2, ‘inflammatory’ DCs, pDCs in the intestinal LP. These findings mirror observations from the SARS epidemic of 2003(*19*), recent autopsy data from COVID-19 patients, and findings from a recently published small animal (golden hamsters) model, all demonstrating a lack of intestinal inflammation despite the presence of viral antigens (*61*).

At a transcriptomic level, our analyses of the intestinal biopsies demonstrated the downregulation of a number of important pro-inflammatory gene products, including *IFNG, IL1B, IKBKB* and *STAT3B*, which contribute to T_H_17 cell differentiation and IBD pathogenesis. Calprotectin, a heterodimer encompassing calgranulin A and calgranulin B, which are encoded by *S100A8* and *S100A9*, respectively, was recently identified as a biomarker of severe COVID-19 disease and suggested to represent a trigger of cytokine release syndrome(*62*). We observed no induction of *S100A8* or *S100A9* in intestinal tissues, even in patients with severe COVID-19, suggesting that the vast intestinal surface did not contribute to the production of calprotectin in COVID-19 patients. Pro-inflammatory cytokines and chemokines that are elevated in the lungs of COVID-19 patients(*63, 64*) were surprisingly downregulated (*IL1B, CXCL8, CXCL2, CXCR6*) or unchanged (*CCL3, CXCL10, CCR1, CXCR3*) in the intestines, compared to non-COVID controls. Other relevant gene products associated with severe COVID-19(*63, 65, 66*) were either significantly lower in the intestinal LP (*CXCL8, IL1B, IFNG, HIF1A, HLA-DQA1*) or trended lower (*CCL2, CCL4, CCL4L2, CTSB, IL23A*) or were comparable to non-COVID controls (*HMGB1, CCL3, CCL8, HMOX1*). Tissue-residence markers such as *ITGA1, CXCR6, JAML*, which are reported to be increased in the pulmonary mucosa of severe COVID-19 patients(*63*), were significantly downregulated in the GI tract. Thus, by dissecting GI tissue transcriptomic response, we noted an immunological milieu characterized by either a suppression of or a failure of induction of an inflammatory response against the virus. Additionally, we observed a lack of production of secondary mediators including calprotectin, which would feed into the inflammatory cascade and culminate in the “cytokine storm” typical of severe COVID-19. Our detailed cellular analyses confirm a lack of inflammatory monocytes and macrophages and a depletion of inflammatory DCs in the GI tract. This failure of induction of inflammatory pathways, regardless of the clinical severity of disease, stands in stark contrast to the massive immunopathology, noted systemically, and within the pulmonary mucosa of severe COVID-19 patients(*67*) as well as patients with active IBD(*68, 69*).

With data demonstrating an attenuated inflammatory response to infection in the GI tract, we sought to address the clinical impact of these findings and recruited two large cohorts from North America and Europe. After controlling for multiple confounding variables including patient age and comorbid illnesses, we found a significant reduction in COVID-19 morbidity and mortality in hospitalized patients in both cohorts. Similar findings were recently reported in a study of 278 patients from a Columbia University Hospital in New York City. Among COVID-19 patients with GI symptoms (diarrhea, nausea and vomiting), there was a non-significant trend towards lower rates of intensive care unit (ICU) admission and a significantly lower rate of death during short-term follow-up(*70*).

In a subset of patients from our large, Discovery and Internal Validation Cohorts, serum samples were available, allowing us to study circulating biomarkers that are associated with COVID-19 severity(*30, 31, 35*). Multiple proinflammatory cytokines/chemokines were found to be downregulated in patients with GI symptoms compared to patients without. These included IL-6 and IL-8, which are now considered a hallmark of increased severity and mortality in COVID-19 as well as a number of other cytokines/chemokines involved in tissue inflammation. The lower IL-17 expression we observed systemically is in line with the reduced T_H_17 RNA seq expression signatures we found in biopsy samples from COVID-19 patients. IL-17 is secreted by T_H_17 and innate cells and is crucial for the recruitment of neutrophils promoting inflammation in a variety of tissues, including the intestinal mucosa(*71, 72*). Additionally, IL-17 has been shown to have pathogenic effects both in SARS-CoV-2(*73*) as well as during SARS-CoV(*74*) and MERS(*75*) infections. Interesting, IL-17C (specifically found to be decreased in patients with GI symptoms in our study) is produced primarily by epithelial cells rather than hematopoietic cells(*76*). Our finding of COVID-19 patients with GI symptoms having reduced CCL28, a cytokine expressed by mucosal epithelial cells and involved in eosinophil chemotaxis(*77*), is consistent with prior work showing increased eosinophils to be associated with severe COVID-19 disease(*31*) and our RNA-seq data showing decreased eosinophil associated genes. IL-15 promotes neutrophilic cytoplasmic re-arrangements and phagocytosis(*78*), DCs differentiation(*79*), and T cell stimulation(*80*). The reduced systemic expression of the IL-15 receptor is in line with our intestinal findings showing lower frequencies of DCs in COVID-19 patients. IL-10 is generally considered an immune modulatory and anti-inflammatory cytokine, however an excess of IL-10 can inhibit the function of immune cells such as NK and CD8+ T cells, possibly delaying clearance of viruses(*81*). Our results of increased systemic expression of IL-10 receptor in patients presenting without GI symptoms is in line with previous studies showing IL-10 to be elevated in COVID-19 patients, associated with disease severity and inversely correlated with CD8^+^ T cells(*82, 83*).

In contrast, the only two cytokines found to be upregulated in patients presenting with GI symptoms were IL-7 and TRAIL, both with important immunoregulatory functions. IL-7, produced by stromal cells and intestinal epithelial cells, has been considered as a therapeutic agent in COVID-19 related to its effects on T cell differentiation and survival(*84*). Similarly, TRAIL is associated with immunosuppressive, immunoregulatory and anti-inflammatory functions(*54, 55*) as well as some evidence of viral infection control in the intestinal tract(*85*).

While our data point to a novel aspect of COVID-19 pathogenesis, we would also like to mention a few limitations of our study. GI biopsies were not performed on a representative subset of the Discovery or Validation Cohorts, but rather from a distinct set of patients who were undergoing clinical indicated procedures. In addition, we recognize our analysis on intestinal mucosal samples were not performed during the most acute phase of the illness in some patients. A separate study by our group that analyzed stool samples from patients going through the acute phase of COVID-19, found little evidence of an active gut inflammatory response. In particular, such stool samples strikingly lacked any significant increase of IL-1*β*, IL-6, TNF-*α* and IL-10, despite the detection of viral genomes (MEDRXIV/2020/183947). Finally, in our clinical outcomes analysis we acknowledge that the reporting GI symptoms can be subject to individual variation and the clinical documentation might vary depending on providers and on the acuity of the patients’ presentation. While it is possible that GI symptoms might have been underreported in patients who were admitted directly to the ICU, our findings are in line with those observed in the External Validation Cohort where patients who died or were admitted to the ICU within 24 hours form initial presentation were excluded from the analysis.

In summary, we have observed an unexpected but significant reduction in COVID-19 severity and mortality when patients demonstrate GI symptoms like diarrhea, nausea or vomiting. These data suggest a previously unappreciated tissue-specific response to SARS-CoV-2 and provide the rationale for more studies aimed at determining the mechanisms underpinning the attenuation of SARS-CoV-2 pathogenicity by the intestinal environment. Such efforts may lead to the development of novel treatments against COVID-19 and potentially other similar deadly infections in the future.

## Materials and Methods

### Clinical cohorts

#### 1. Intestinal Biopsy Cohort

Intestinal biopsies and peripheral blood from 18 COVID-19 and 10 control patients were obtained between April 17, 2020 and June 2, 2020. Subjects included hospitalized patients as well as those seen in the outpatient GI practices. COVID-19 cases and controls were defined on the basis of nasopharyngeal SARS-CoV-2 swab PCR tests. The demographic characteristics of these patients and controls are provided in Fig. 1A, Table 1, and table S1. Informed consent was obtained from all patients. The biopsy-related studies were approved by the Mount Sinai Ethics Committee/IRB (IRB 16-0583, The impact of viral infections and their treatment on gastrointestinal immune cells). COVID-19 severity was defined based on internal scoring system developed by the Department of Infectious Diseases at Mount Sinai Hospital. This scoring system was developed according to the WHO Ordinal Clinical Progression/Improvement Scale (https://www.who.int/publications/i/item/covid-19-therapeutic-trial-synopsis) and based on oxygenation status and organ damage, with the following definitions: Mild - SpO2>94% on room air AND no pneumonia on imaging, Moderate - SpO2<94% on room air OR pneumonia on imaging, Severe - high flow nasal cannula (HFNC), non-rebreather mask (NRBM), Bilevel Positive Airway Pressure (non-invasive positive airway ventilation), or Mechanical ventilation AND no pressor medications AND creatinine clearance > 30 AND ALT < 5x upper limit of normal, Severe with evidence of end organ damage (EOD) - high flow nasal canula (HFNC), non-rebreather mask (NRBM), Bilevel Positive Airway Pressure (non-invasive positive airway ventilation), or Mechanical ventilation AND pressor medications OR creatinine clearance <30 OR new renal replacement therapy OR ALT > 5x upper limit of normal (table S2).

#### 2. Discovery Cohort

Patients admitted to Mount Sinai Hospital (MSH) between April 1, 2020 and April 15, 2020 were recruited into the Discovery Cohort if they were SARS-CoV-2 PCR positive, more than 18 years of age and if the “ELLA panel of cytokines” (IL-6, IL-8, IL-1*β* and TNF-*α*) was performed as part of clinical care. Clinical details from eligible patients were extracted from Mount Sinai Data Warehouse (MSDW) under an IRB approved protocol (IRB-20-03297A North American registry of the digestive manifestations of COVID-19).

A total of 634 subjects were included in the Discovery Cohort (fig. S11). In addition to demographic information (including race and ethnicity and primary language), clinical characteristics, laboratory data and outcomes data was extracted from the medical charts. Co-variates that were studied included: history of smoking, BMI (obesity defined as BMI >30) and comorbid conditions including, hypertension, diabetes, chronic lung disease (including asthma and COPD), heart disease (including coronary artery disease, atrial fibrillation and heart failure), chronic kidney disease, cancer, HIV, and inflammatory bowel disease (IBD).

GI symptoms were defined as more than one episode of either diarrhea, nausea, and/or vomiting at the time of admission. If only one episode of either diarrhea, nausea, and/or vomiting was specifically documented, patients were not considered to have GI symptoms. Additionally, we did not consider GI symptoms that developed during the course of hospitalization, as they could reflect nosocomial or treatment-related effects and only considered the GI symptoms that were present at the time of hospital admission so as to avoid including iatrogenic confounders (treatments or hospital acquired illnesses that can result in diarrhea, nausea and vomiting).

Disease severity (as described above) and mortality were considered as outcomes variables. Mortality was calculated as patient status (dead or alive) at 25 days post admission. If no information was available after discharge, patients were censored at the time of hospital discharge.

#### 3. External Validation Cohort

To confirm the Discovery Cohort findings, we analyzed a cohort of patients admitted to a tertiary care center in Milan, Italy between February 22, 2020 and March 30, 2020. A total of 287 patients with a confirmed positive SARS-CoV-2 PCR and who did not die or were not transferred to the ICU within 24 hours from admission were studied. Presence of vomiting and diarrhea (defined as at least three loose bowel movement per day) on or prior to admission was recorded. Outcomes were analyzed using ICU admission, death or the composite study end-point of ICU admission or death within 20 days of hospitalization.

#### 4. Internal Validation Cohort

To test a predictive model of COVID-19 severity and disease-related mortality, we developed a distinct ‘Internal Validation Cohort’ of patients who were hospitalized at MSH between April 16, 2020 and April 30, 2020 and satisfied the same inclusion and exclusion criteria as in the Discovery Cohort. Additionally, patients already included in the Discovery Cohort were excluded. From a total of 408 patients, 242 met inclusion criteria and were thus included in the Internal Validation Cohort. Demographic, clinical and outcomes related data was extracted from patients’ medical records as described for the Discovery Cohort.

### SARS-CoV-2 testing

The SARS-CoV-2 PCR was run in the Clinical Microbiology laboratory as part of routine care on the Roche cobas platform. This platform performs selective amplification of 2 targets ORF-1 gene (Target 1) and the E-gene for pan-Sarbecovirus (Target 2) (detects SARS-CoV-2 as well as SARS or MERS viruses, but not routine seasonal Coronavirus). A positive result indicated that either both Target 1 and Target 2 were detected (majority of cases) or Target 1 alone was detected. A presumptive positive result indicates a negative Target 1 result and a positive Target 2 result which according to the manufacture can be a result of the following: “1) a sample at concentrations near or below the limit of detection of the test, 2) a mutation in the Target 1 target region in the oligo binding sites, or 3) infection with some other Sarbecovirus (e.g., SARS-CoV or some other Sarbecovirus previously unknown to infect humans), or 4) other factors.” Patients with a presumptive positive SARS-CoV-2 PCR were included in the analysis if they were treated clinically as having COVID-19.

### Immunofluorescent (IF) microscopy

Formalin fixed, paraffin embedded tissue acquired during routine clinical care was obtained from the pathology core at our institution. Sections (5µm) were dewaxed in xylene and rehydrated in graded alcohol and then washed in phosphate-buffered saline (PBS). Heat-induced epitope retrieval was performed by incubating slides in a pressure cooker for 15 minutes on high in target retrieval solution (Dako, S1699). Slides were then left to cool in the solution at room temperature for 30 minutes. Slides were washed twice in PBS and then permeabilized for 30 minutes in 0.1% tritonX-100 in PBS. Non-specific binding was blocked with 10% goat serum for 1 hour at room temperature. Sections were then incubated in primary antibodies diluted in blocking solution overnight at 4°C. Primary and secondary antibodies are summarized in table S15. Slides were washed in PBST (0.1% tween 20, PBS) thrice and then incubated in secondary antibody and 4′,6-diamidino-2-phenylindole (1μg/mL) for 1 hour at room temperature. Sections were washed twice in PBST and once in PBS then mounted with Fluoromount-G (Electron microscopy sciences, 1798425). Controls included, omitting primary antibody (no primary control), or substituting primary antibodies with non-reactive antibodies of the same isotype (isotype control). Tissue was visualized and imaged using a Nikon Eclipse Ni microscope and digital SLR camera (Nikon, DS-Qi2).

### Electron Microscopy (EM)

Biopsy specimens for electron microscopy were placed in 3% buffered glutaraldehyde. Following post-fixation in 1% osmium tetroxide, tissues were serially dehydrated and embedded in epoxy resin in standard fashion. One-micron toluidine-stained scout sections were prepared for light microscopic orientation; 80nm ultrathin sections for EM were stained with uranyl acetate and lead citrate and examined in a Hitachi 7650 transmission electron microscope at 80kV.

### Biopsy collection and processing for Mass cytometry (CyTOF)

Endoscopic biopsies were obtained from COVID-19 patients and controls during clinically indicated endoscopic procedures. The biopsies were processed in biosafety level 3 (BSL-3) facility within 2 hours of collection.

Briefly, biopsies were transferred to 10 ml of ‘dissociation buffer’ (1M HEPES(Lonza), 5μM EDTA(Invitrogen), 10% FBS in HBSS buffer (Gibco). The tubes were kept in a shaker (180 rpm, 37°C) for 20 min and then gently vortexed. Cell suspensions were collected after passing the biopsies through 100μm cell strainers. A second round of EDTA dissociation was performed as detailed above. The cell suspension was centrifuged at 1800 rpm to pellet the epithelial fraction and kept on ice. The remaining tissue was transferred to fresh tubes containing a ‘digestion buffer’ (2% FBS, 0.005g Collagenase type IV per sample (Sigma), 100 μl DNAse-I (Sigma) in RPMI). Tubes were placed in the shaker (180 rpm, 37°C) for 40 min and thereafter gently vortexed. The digested tissues were filtered through 100 μm cell strainers followed by another round of filtration through 40μm cell strainers. Cell suspensions were centrifuged at 1800 rpm to obtain lamina propria mononuclear cells. Both epithelial cell (EC) and lamina propria (LP) pellets were then resuspended into 500 μl of RPMI (Gibco) containing 10% FBS+ 1μl Rh103 +1μl IdU and incubated at 37°C for 20 min. 5 ml RPMI (+10%FBS) was added to each tube and spun at 1800 rpm to pellet cells. 700 μl of Prot1 stabilizer (SmartTube Inc.) was added to each tube and transferred to cryovials and incubated at room temperature for 10 min. Cryovials were immediately transferred to -80°C until the sample was acquired for mass cytometry as detailed below.

### Blood collection and processing for CyTOF

Phlebotomy was performed on COVID-19 patients and non-COVID-19 controls at the time of endoscopic evaluation. All the blood samples from COVID-19 patients were processed in enhanced BSL2 conditions as per institutional guidelines. Briefly, 15ml of Lymphosep*®* - Lymphocyte Separation Medium (MP Bio.) was added to each 50 ml centrifugation tube. Blood was diluted with PBS to bring the volume up to 30ml and diluted blood was layered gently over Lymphosep*®*. Tubes were then centrifuged at 2000 rpm for 20 mins with the brakes and acceleration off. After centrifugation, the buffy coat containing PBMCs was transferred to another tube and was centrifuged at 1800 rpm to pellet the cells. Pellets were resuspended in PBS and tubes were centrifuged at 1800 rpm. Finally, the pellets were resuspended in the freezing medium (10% DMSO + 44% FBS in RPMI) and cryopreserved at -80 °C.

### CyTOF processing and data acquisition

Cells were processed as previously described by *Geanon et al*.(*86*). Briefly, EC and LP SmartTube proteomic stabilized samples were thawed in a 10°C water bath and washed with Cell Staining Buffer (Fluidigm). To facilitate data acquisition and doublet removal, multiple samples were also barcoded using Fluidigm Pd barcoding kits and then washed and pooled for data acquisition. Immediately prior to data acquisition, samples were washed with Cell Staining Buffer and Cell Acquisition Solution (Fluidigm) and resuspended at a concentration of 1 million cells per ml in Cell Acquisition Solution containing a 1:20 dilution of EQ Normalization beads (Fluidigm). The samples were then acquired on a Helios Mass Cytometer equipped with a wide-bore sample injector at an event rate of <400 events per second. After acquisition, repeat acquisitions of the same sample concatenated and normalized using the Fluidigm software, and barcoded samples were de-multiplexed using the Zunder single cell debarcoder.

### CyTOF Data analysis

De-barcoded files were uploaded to Cytobank for analyses. Immune cells were identified based on Ir-193 DNA intensity and CD45 expression; Ce140+ normalization beads, CD45-low/Ir-193-low debris and cross-sample and Gaussian ion-cloud multiplets were excluded from subsequent downstream analysis. Major immune cell types were identified using automated Astrolabe approach, the result of which largely correlated well with our manual gating approaches. The impact of each tested condition on relative staining quality was evaluated in two ways: 1) overall correlations were determined by calculating the Pearson’s correlation coefficients for the median expression of each marker across each defined immune subset; and 2) a staining index was calculated using defined populations showing the highest and lowest expression levels of each marker: SI = (Medianpos - Medianneg) / 2 X Std.Devneg. It is already been described that SmartTube-based fixation protocols take into account previously described mass cytometry artifacts such as cell-cell multiplets, isotopic spillover or oxidation, or mass cytometer instrument configuration(*86*).

### Statistical Analysis for CyTOF

Pre-gated viable CD45+ cells were first clustered and annotated using the Astrolabe Cytometry Platform (Astrolabe Diagnostics, Inc.), which involves using a hierarchy-based FlowSOM algorithm for labeling cell populations in individual samples. These Astrolabe Profiling clusters from each tissue type were then meta-clustered across all samples utilizing Clustergrammer2’s interactive heatmap as a method to interrogate antibody expression across every cluster and curate and assign cell population categories. Single sample clusters were also visualized using UMAP. Pairwise comparisons were performed on the frequencies of each identified cell population between the patient cohorts (COVID-19 vs. control, COVID-19 severe vs. control, COVID19-asymptomatic/mild/moderate vs. control) to determine fold change, p-values and FDR adjusted p-values using the Benjamini-Hochberg(*87*) method to account for multiple comparisons.

### Cell Culture Experiments and Virus Isolation

African green monkey kidney epithelial cells (Vero E6) were originally purchased from American Type Culture Collection (ATCC). Cells were maintained in Dulbecco’s modified Eagle’s medium (DMEM) w/ L-glutamate, sodium pyruvate (Corning) supplemented with 10% fetal bovine serum (FBS), 100 U penicillin/ml, and 100 mg streptomycin/ml. For all experiments, the cells were always maintained in monolayers.

Several attempts were made to isolate live infectious particles from these biopsies. Briefly, biopsies were collected and stored in PBS until homogenization. Following homogenization and centrifugation (10,000 × g, 20 min, 4°C), the resulting supernatant tissue supernatant was inoculated onto Vero E6 monolayer maintained in optimal virus growth media for SARS-CoV-2 virus (DMEM w/ L-Glutamate, Sodium Pyruvate, 2% FBS, 100 U Penicillin/ml, and 100 mg Streptomycin/ml, 10 mM Non-Essential Amino Acids, 1 mM Sodium Pyruvate and 10 mM HEPES). Vero E6 cells were incubated at 37 °C, 5% CO2 for a week and monitored daily for potential cytopathic effect (CPE).

Cell culture supernatants were also collected and assessed for the presence of infective particles by plaque assay. Briefly, ten-fold serial dilutions were performed in infection media for SARS-CoV-2 and inoculated onto confluent Vero E6 cell monolayer in 6-well plate. After one-hour adsorption, supernatants were removed, and cells monolayers were overlaid with minimum essential media (MEM) containing 2% FBS and purified agar (OXOID) at a final concentration of 0.7%. Cells were then incubated for 3 days at 37°C. Cells were fixed overnight with 10% formaldehyde for the inactivation of potential SARS-CoV-2 virus. Overlay was removed and cells were washed once with PBS. A 2% crystal violet solution was used for plaque visualization and count. Experiments were performed under BSL3 conditions.

### Specimen Processing for Nucleic Acid Extraction

Whole biopsy tissues from COVID-19 patients and non COVID controls were directly homogenized in Trizol (Invitrogen) and used to detect the presence of viral RNA. Total RNA was extracted using Direct-zol RNA Miniprep Plus (Zymo) kit according to the manufacturer’s instructions. In parallel, RNA isolated from both the intestinal compartments, EC and LP cellular fractions, were used for transcriptomics analyses.

### RT-qPCR for Viral Determination

RNA was retro-transcribed using the enzyme Maxima First Strand cDNA (Thermofisher), and PCR reaction was performed using the TaqMan™ Universal PCR Master Mix (Thermofisher). For detection of SARS-CoV-2 RNA, we used the following primers and probe targeting the N gene: N1-SARS-CoV2-F5’GACCCCAAAATCAGCGAAAT;

N1-SARS-CoV2-R5’TCTGGTTACTGCCAGTTGAATCTG; N1-SARS-CoV2-P FAM5’ ACCCCGCATTACGTTTGGTGGACC-BHQ-1 (https://www.cdc.gov/coronavirus/2019-ncov/lab/rt-pcr-panel-primer-probes.html) Another set of primers was included to detect host 18S and GAPDH. Limit of detection and amplification efficiency were calculated before GI tissue quantification for N1 (173%; R^2^>0.94) primers using plasmids expressing the NP protein (2019-nCoV_N_Positive Control, IDT, cat. 10006625). RNA from GI tissues were run in triplicates in 384-well plates using the following cycling conditions on the Roche LightCycler 480 Instrument II (Roche Molecular Systems, 05015243001): 50°C for 2 min; 95°C for 10 min; 40 cycles of 95°C for 15 sec and 60°C for 1 min. Samples below the limit of detection (C_t_≥34 corresponding to 100 genome copies/reaction) were considered negative. Primers and probes for housekeeping genes GAPDH (assay ID Hs02758991_g1 FAM) and 18S (assay ID Hs03928990_g1 FAM) were obtained from Thermofisher Scientific).

### RNA Sequencing

#### Library preparation and sequencing

RNA-sequencing (RNA-seq) was performed on RNA isolated from the EC and LP samples obtained from COVID-19 cases and controls. Directional RNA-seq libraries were prepared from 50 ng of total RNA with the TruSeq® Stranded Total RNA prep with Ribo-Zero kit (Cat no. 20020599). Paired-end (100 bp) sequencing was performed for DNA libraries on an Illumina NovaSeq instrument on a NovaSeq S1 Flowcell, with an average yield of 39 million PE reads/sample.

#### RNA-seq analysis

Base-calling and quality scoring of sequencing data were done through Illumina’s Real-Time Analysis (RTA) software. RNA-seq data processing and reference mapping were done with custom analysis scripts combining publicly available tools as previously described(*88*) with modifications as follows, reads were mapped to a custom reference that combined the human hg38 reference genome (Release 34, GRCh38.p13) and the SARS-CoV-2 genome (RefSeq NC_045512) for simultaneous quantification of host and virus transcripts.

Differential gene expression (DGE) analysis was performed with the Bioconductor edgeR package (*89*) using as input a combined matrix of mapped paired-end read raw counts, with genes in rows and samples in columns. Prior to DGE analysis, gene counts were converted to fragments per kb per million reads (FPKM) with the RSEM package with default settings in strand-specific mode(*90*).

Genes with less than 1 FPKM in at least 50% of the samples were removed. The remaining gene counts were then normalized across samples using the weighted trimmed mean of M-values (TMM) method(*91*). The dispersion was estimated by fitting a generalized linear model (GLM) as implemented in edgeR, sex was fitted as a covariate on a per-patient paired design. Pairwise comparisons were performed between sample groups (i.e., between tissue sections, and between cases and controls). Significant expression differences were selected based on eBayes adjusted p values corrected for multiple testing using the Benjamini-Hochberg method (q ≤ 0.05).

#### Gene Ontology and Pathway Enrichment Analysis

KEGG pathway and gene ontology (GO) biological process (BP), molecular function (MF), and/or cellular component (CC) enrichment analyses were performed using the gProfileR R v0.6.8 package(*92*). The background gene set was restricted genes with detected expression (defined as genes with expression levels above 1 FPKM in at least 50% of samples). Genes with differential expression were ranked by log 2 fold change and used as an ordered query. P values were corrected using the g:SCS algorithm to account for multiple comparisons.

#### Cell-type deconvolution and gene signature enrichment analysis

For cell-type deconvolution of the bulk RNA-seq data, Gene Set Enrichment Analysis (GSEA) of differentially expressed genes of cases vs controls comparisons was performed against cell type gene-expression single-cell signatures from intestinal mucosa(*44*) and gene-expression signatures from ileal dendritic cell (DCs) subsets (*43*). Similarly, differentially expressed genes were tested for enrichment of gene signatures associated with an antiviral response, inflammation, and cytokine signaling in acutely infected post-mortem tissue with SARS-CoV-2(*30*), were tested for significant (p ≤ 0.05) enrichment using Fisher’s exact tests and using Bonferroni correction for multiple comparisons.

Additionally, GSEA(*93*) was carried out on a rank ordered list of the infected EC versus control molecular analysis. The ranking metric used was logFC * -logP value, however, the results were similar when logFC metric was also used (data not shown). For the COVID-19 associated datasets, we curated two signatures from infected organoids(*40*): hSIOs-COVID-19: human small intestinal organoids (hSIOs) grown in either i) Wnt high expansion (EXP) medium (at adjP<0.05) or ii) differentiation (DIF) medium (at adjP<0.1). The standard GSEA settings were used, namely ‘meandiv’ for normalization mode, ‘weighted’ enrichment statistic, and ‘1000’ permutations. GSEA using the Hallmark database (v7.1, (*94*)) was also performed with the same settings.

### Computational analyses

#### Descriptive statistics

Basic demographics and clinical characteristics of the cohort were defined by descriptive statistics. For univariable statistical analyses, graph pad prism (version 8) was used. For age, an unpaired two tailed t-test was performed. For categorical variables, the Fisher’s exact test or the Chi-square test was used as appropriate.

#### Multivariate model based on Discovery Cohort

For this analysis, we considered 570 patients with clinical descriptors including as age, gender, race, BMI, comorbidities and GI symptoms. A multivariate logistic regression was utilized to model severity and mortality as function of each of the GI symptoms and clinical variables including race, age, gender, BMI, heart and lung diseases and hypertension. In particular, race was stratified as White (Caucasian), Black (African-American), Hispanic and others; lung disease was set equal to 1 if the patient was either affected by COPD or asthma and zero otherwise; heart disease was set equal to 1 if the patient was either affected by coronary artery disease, atrial fibrillation or heart failure and 0 otherwise. The severity indicator was set equal to 1 for severe and severe with EOD patients and 0 for Mild and Moderate COVID patients; mortality was set equal to 1 for deceased patients and 0 otherwise. Significant association based on 95% confidence interval (CI) are reported in table S9. CI of odds ratio were computed based on 1000 bootstrap iterations. At each bootstrap iteration, patients were sampled with replacements and logistic regressions were estimated considering as outcome severity and mortality. Then, 95% CI of coefficients and odds ratio were estimated across bootstrap iterations (Fig. 5B).

#### External Validation Cohort

For this analysis, we considered 228 patients with clinical data such as age, gender and GI symptoms as described in *Aghemo et al*.(*95*). A multivariate logistic regression was utilized to model mortality, ICU admission and the composite outcome of ICU admission or mortality as function of presence or absence of diarrhea and clinical variables including age, gender, BMI, heart disease, COPD, diabetes and hypertension. Heart disease was set equal to 1 if the patient was either affected by coronary artery disease or atrial fibrillation and 0 otherwise. Confidence intervals of odds ratio were computed based on 1000 bootstrap iterations. At each bootstrap iteration, patients were sampled with replacements and logistic regressions were estimated considering the outcome as mortality, ICU admission or the composite outcome of ICU admission or death. Then, 95% confidence intervals of odds ratio were estimated across bootstrap iterations (Fig. 5C).

#### Predictive performance based on the Internal Validation Cohort

For this analysis, we considered 233 patients with clinical data including age, BMI, and GI symptoms. In order to evaluate the predictive performance of each model, bootstrapping was performed. Specifically, at each bootstrap iteration, we randomly sampled patients in the Discovery Cohort with replacement and estimated a logistic regression to model each outcome as function of a particular GI symptom, age and BMI. In this analysis, only age and BMI were adjusted for since they were the only variables significantly associated with both outcomes across different GI symptoms models in the Discovery Cohort (Fig. 5B). Then, the estimated model was utilized to predict the outcome of patients in the Internal Validation Cohort. This procedure was repeated for 1000 bootstrap iterations. For each iteration, Receiving Operating Characteristic (ROC) curve and area under the curve (AUC) were computed. For comparison purposes, the distribution of AUC across 1000 bootstrap iterations from the predictive model based on age and BMI only was considered. Fig. 5D shows the boxplot of AUC values across 1000 bootstrap iterations. Then, considering the following model

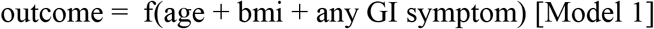

we evaluated the effect of each variable on the outcome by computing the reduction in AUC obtained after removing one variable at a time. For this purpose, the AUC of model [Model 1] was compared to the following three models

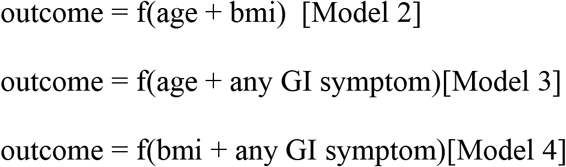

for 1000 bootstrap iterations. Following the strategy above, at each bootstrap iteration, patients were sampled with replacement. Fig. 5E shows the 95% confidence intervals of difference in AUC between [Model 1] and [Model 2], [Model 3] and [Model 4] (i.e., AUCModel1 - AUCModel2, AUCModel1 - AUCModel3, AUCModel1 - AUCModel4) across 1000 bootstrap iterations. The difference in AUC was computed considering both mortality and severity as the outcome.

### Quantification of SARS-CoV-2 nasopharyngeal viral loads

SARS-CoV-2 viral loads were determined as detailed in Pujadas et al(*53*). Briefly, viral RNA was extracted from the nasopharyngeal swab specimen followed by real time RT-PCR using N2 primers. Only specimens with N2_Cpt_ < 38 were included. SARS-CoV-2 viral RNA was calculated with the delta CT method and a standard curve. Viral loads are presented as log base 10 transformed uncorrected N2 value + 1000 (constant added before transformation)(*53*). For patients with multiple NP swabs available, the first swab was used for analysis.

### ELLA Cytokine panel

The ELLA platform is a method for rapid cytokine measurement using microfluidics ELISA assays. The assay measured TNF-α, IL-6, IL-8, and IL-1β, previously validated by the Mount Sinai Human Immune Monitoring Center (HIMC) using plasma from multiple myeloma patients and recently reported for large cohort of COVID-19 patients admitted to Mount Sinai Hospital(*35*).

### Multiplexed proteomic assay (Olink)

For analysis of circulating cytokines, we used a multiplexed proteomic inflammation panel (Olink), which consists of 92 inflammation-related proteins quantified by an antibody-mediated proximity extension-based assay(*79*). Samples with normalized protein expression values below the limit-of-detection in >75% of samples were excluded from further analysis. For the remainder of analytes, any sample under the limit of detection was assigned a value of the limit-of-detectionv divided by the square root of 2. The log2 fold-change over the median healthy control protein expression was then calculated, and the Benjamini-Hochberg(*80*) procedure was used to adjust P values for multiple testing.

### Consensus Clustering of Olink Data

For this analysis, we considered 238 samples with GI symptoms annotation. Consensus clustering was performed based on the abundance of 92 cytokines across all 238 samples. Consensus clustering was performed using the R packages ConsensusClusterPlus(*81*) based on z-score normalized data. Specifically, markers were partitioned into six clusters using the Kaplan Meier (KM) algorithm, which was repeated 1000 times. Then, markers in each cluster were considered in order to derive cluster z-score signatures via package GSVA(*82*). Based on these signatures, the association between different clusters and GI symptoms were derived via logistic regression with outcome corresponding to each GI symptom. Fig. 6C shows the signed FDR (-log10 scale). P-values were adjusted via Benjamini-Hochberg(*80*).

### Defining associations between GI symptoms and Olink protein markers

Associations between GI symptoms and Olink proteomic data were derived using unpaired t-test comparing the symptomatic and asymptomatic groups. P-values were adjusted via Benjamini-Hochberg(*80*). Only associations passing a 10% FDR were reported as significant (Fig. 6D).

### Defining associations between GI symptoms and ELLA cytokine markers

Unpaired two-tailed t-tests were used to compare individual cytokines quantified by the ELLA panel between GI symptomatic and asymptomatic groups. P-values were adjusted via Benjamini-Hochberg(*80*).

## Supplementary Materials

Fig. S1. Sample allocation for different assays in COVID-19 patients and controls.

Fig. S2. Representative H&E staining of small intestinal biopsies of COVID-19 patients.

Fig. S3. Representative IF of small intestinal biopsies of COVID-19 patients.

Fig. S4. Representative IF of small intestinal biopsies of control patients.

Fig. S5. Altered immune populations in the lamina propria of COVID-19 patients compared to controls.

Fig. S6. Altered immune populations in the epithelial compartment (EC) of COVID-19 patients compared to controls.

Fig. S7. Altered immune populations in the blood of COVID-19 patients compared to controls.

Fig. S8. Altered T cell populations in blood and intestinal biopsies of COVID-19 patients compared to controls based on manual gating of the populations.

Fig. S9. Distinct expression profiles in the intestinal epithelial compartment (EC) and lamina propria (LP).

Fig. S10. Immune signature in the epithelial fraction of COVID-19 patients. Fig. S11. Flow diagram of the Discovery Cohort.

Fig. S12. GI symptoms associated with reduced mortality and severity.

Fig. S13. COVID-19 patients with GI symptoms have reduced levels of circulating IL-6 and IL-8.

Fig. S14. Correlation matrix (Pearson’s) for 92 markers contained in the Olink platform. Table S1. Clinical characteristics of SARS-CoV-2 uninfected control patients who underwent endoscopic biopsies of the GI tract.

Table S2. Criteria for scoring disease severity in COVID-19 patients.

Table S3. Clinical characteristics of COVID-19 patients including their symptoms on presentation, co-morbidities and treatment regimens.

Table S4. Histopathological characteristics of COVID-19 patients. Table S5. Histopathological characteristics of pre-pandemic controls.

Table S6. Discovery Cohort basic demographics, clinical characteristics and outcomes.

Table S7. COVID-19 disease severity and mortality in patients with and without GI symptoms in the Discovery Cohort.

Table S8. Basic demographics in survivors and non-survivors in the Discovery Cohort. Table S9. Confidence intervals of odds ratio based on 1000 bootstrap iterations for severity, mortality and ICU admission in the Discovery Cohort.

Table S10. Age, gender and mortality in an External Validation (Italian) Cohort stratified by presence or absence of diarrhea on admission.

Table S11. Confidence interval of AUC (95%) based on 1000 bootstrap iterations for severity, mortality and ICU admission in the Internal Validation Cohort.

Table S12. IL-6, IL-8, TNF-α, and IL-1β concentrations on admission in patients with and without GI symptoms.

Table S13. Cluster assignment for each of the 92 Olink analytes.

Table S14. Olink analytes in patients with and without GI symptoms. Table S15. List of antibodies used for microscopy studies.

Data file S1. Antibody panel used for mas cytometry or CyTOF analyses (provided as separate Excel file).

Data file S2. Frequencies of different immune populations in intestinal biopsies (lamina propria and epithelial compartment) and in blood in COVID-19 patients and controls (provided as separate Excel file).

Data file S3. RNA sequencing table (provided as separate Excel file).

## Data Availability

Mass cytometry and RNA sequencing data are provided in the supplementary information.

## Acknowledgements

We would like to thank the clinical staff, physicians and patients who participated in this study. This research was partly funded by NIH/NIDDK123749 (SM). Additional support was provided by CRIP (Center for Research for Influenza Pathogenesis), a NIAID supported Center of Excellence for Influenza Research and Surveillance (CEIRS, contract # HHSN272201400008C), and NIAID R01AI113186 (to H.B). Additionally, the work was supported by the generous support of the JPB Foundation, the Open Philanthropy Project (research grant 2020-215611 (5384)), the Defense Advanced Research Projects Agency, and anonymous donors to AG-S. MT was funded by the Digestive Disease Research Foundation (DDRF). A.S.G-R. is supported in part by a Robin Chemers Neustein Postdoctoral Fellowship Award. The research carried out by H.V.B and A.S.G-R was supported by the Office of Research Infrastructure of the National Institutes of Health (NIH) under awards S10OD018522 and S10OD026880. S.T.C. is supported by grant F30CA243210. G.J.B. is supported by a Research Fellowship Award from the Crohn’s and Colitis Foundation of America. M.P.S. is supported by NIH T32 5T32AI007605. S.G. is supported by grants U24 CA224319, U01 DK124165, and P01 CA190174. We also thank Randy Albrecht for support with the BSL3 facility and procedures at the Icahn School of Medicine at Mount Sinai (ISMMS).

## Authors’ abbreviations

A.E.L.: Gastroenterology Fellow, D.J.: Postdoctoral Fellow, F.C.: Assistant Professor, A.S.G-R.: Postdoctoral Fellow, M.T.: MD candidate, T.A.: Instructor, T.L.P.: Gastroenterology Fellow, I.R.: Assistant Professor, K.D.: Internal Medicine Resident, B.L.: Computational Scientist, R.D.: Senior Research Coordinator, S.T.C.: MD/PhD candidate, G.M.D.: Research manager, S.N.: Associate Professor, H.M.K.: Assistant Professor, B.S.G.: Assistant Professor, G.N.: Assistant Professor, E.P: Pathology Resident, J.R.: Electron Microscopist, S.N.: Assistant Professor, A.G.: Assistant Professor, J.A.: Professor, M.T.: Research Coordinator, R.G.: Research Professor, K.S.: Master’s candidate, J.H: Associate Professor, G.J.B.: Instructor, A.C.L.: MD/PhD candidate, M.P.S.: MD/PhD candidate, T.P.: PhD candidate, P.W.: Professor, A.C.: Professor, J.J.F.: Associate Professor, J.F.C.: Professor, E.K.: Assistant Professor, C.A.: Associate Professor, M.M.: Professor, S.G.: Associate Professor, N.H.: Professor, S.D.: Professor, C.C.C.: Professor, A.R.: Associate Professor, N.A.K.: Associate Professor, A.A.: Associate Professor, F.P.: Assistant Professor, H.V.B.: Assistant Professor, A.G.S.: Professor, S.M.: Associate Professor.

## Data and materials availability

Will be made available upon acceptance.

