## Supplemental Materials for "Gastrointestinal involvement attenuates COVID-19 severity and mortality"

**Supplementary Materials**

Fig. S1. Sample allocation for different assays in COVID-19 patients and controls.

Fig. S2. Representative H&E staining of small intestinal biopsies of COVID-19 patients.

Fig. S3. Representative IF of small intestinal biopsies of COVID-19 patients.

Fig. S4. Representative IF of small intestinal biopsies of control patients.

Fig. S5. Altered immune populations in the lamina propria of COVID-19 patients compared to controls.

Fig. S6. Altered immune populations in the epithelial compartment (EC) of COVID-19 patients compared to controls.

Fig. S7. Altered immune populations in the blood of COVID-19 patients compared to controls.

Fig. S8. Altered T cell populations in blood and intestinal biopsies of COVID-19 patients compared to controls based on manual gating of the populations.

Fig. S9. Distinct expression profiles in the intestinal epithelial compartment (EC) and lamina propria (LP).

Fig. S10. Immune signature in the epithelial fraction of COVID-19 patients.

Fig. S11. Flow diagram of the Discovery Cohort.

Fig. S12. GI symptoms associated with reduced mortality and severity.

Fig. S13. COVID-19 patients with GI symptoms have reduced levels of circulating IL-6 and IL-8.

Fig. S14. Correlation matrix (Pearson's) for 92 markers contained in the Olink platform.

Table S1. Clinical characteristics of SARS-CoV-2 uninfected control patients who underwent endoscopic biopsies of the GI tract.

Table S2. Criteria for scoring disease severity in COVID-19 patients.

Table S3. Clinical characteristics of COVID-19 patients including their symptoms on presentation, co-morbidities and treatment regimens.

Table S4. Histopathological characteristics of COVID-19 patients.

Table S5. Histopathological characteristics of pre-pandemic controls.

Table S6. Discovery Cohort basic demographics, clinical characteristics and outcomes.

Table S7. COVID-19 disease severity and mortality in patients with and without GI symptoms in the Discovery Cohort.

Table S8. Basic demographics in survivors and non-survivors in the Discovery Cohort.

Table S9. Confidence intervals of odds ratio based on 1000 bootstrap iterations for severity, mortality and ICU admission in the Discovery Cohort.

Table S10. Age, gender and mortality in an External Validation (Italian) Cohort stratified by presence or absence of diarrhea on admission.

Table S11. Confidence interval of AUC (95%) based on 1000 bootstrap iterations for severity, mortality and ICU admission in the Internal Validation Cohort.

Table S12. IL-6, IL-8, TNF-α, and IL-1β concentrations on admission in patients with and without GI symptoms.

Table S13. Cluster assignment for each of the 92 Olink analytes.

Table S14. Olink analytes in patients with and without GI symptoms.

Table S15. List of antibodies used for microscopy studies.

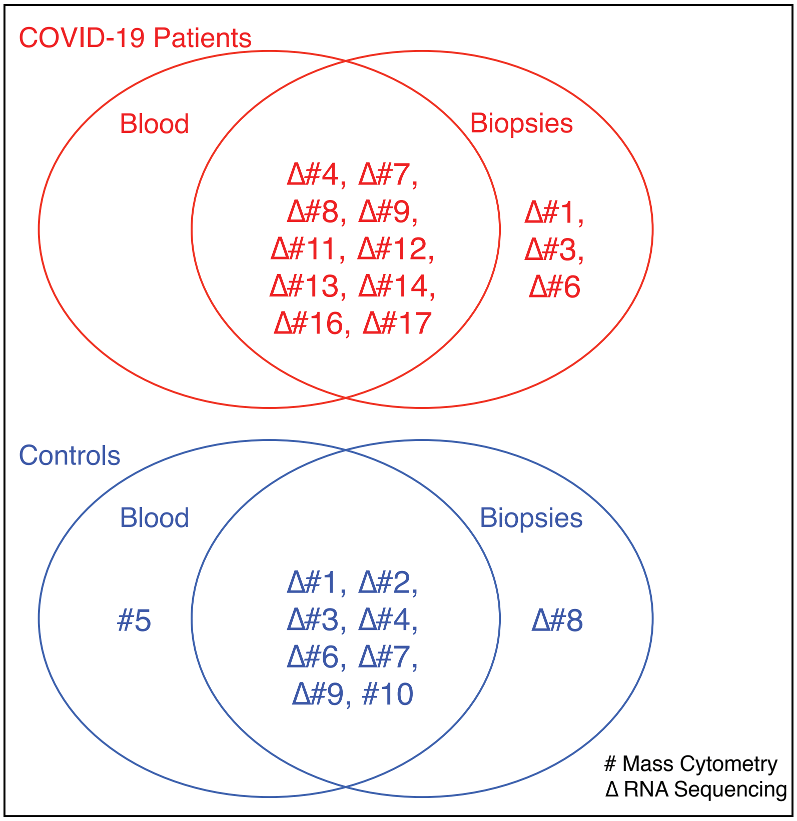

| Assays | Sample | COVID-19 cases | Controls |
|---|---|---|---|
| **Mass Cytometry** | Biopsy | 13 | 9 |
| | Blood | 10 | 9 |
| **RNA Sequencing** | Biopsy | 13 | 8 |
| | Blood | N/A | N/A |

**Fig. S1 Sample allocation for different assays in COVID-19 patients and controls.** Venn diagrams showing blood and biopsy samples used for mass cytometry (#) and RNA sequencing (Δ) in COVID-19 patients (red) and controls (blue). The numbers in the Venn diagrams refer to respective patient and control cases in Table 1 and table S1. The table summarizes the total number of blood and biopsy samples allocated for mass cytometry and RNA sequencing.

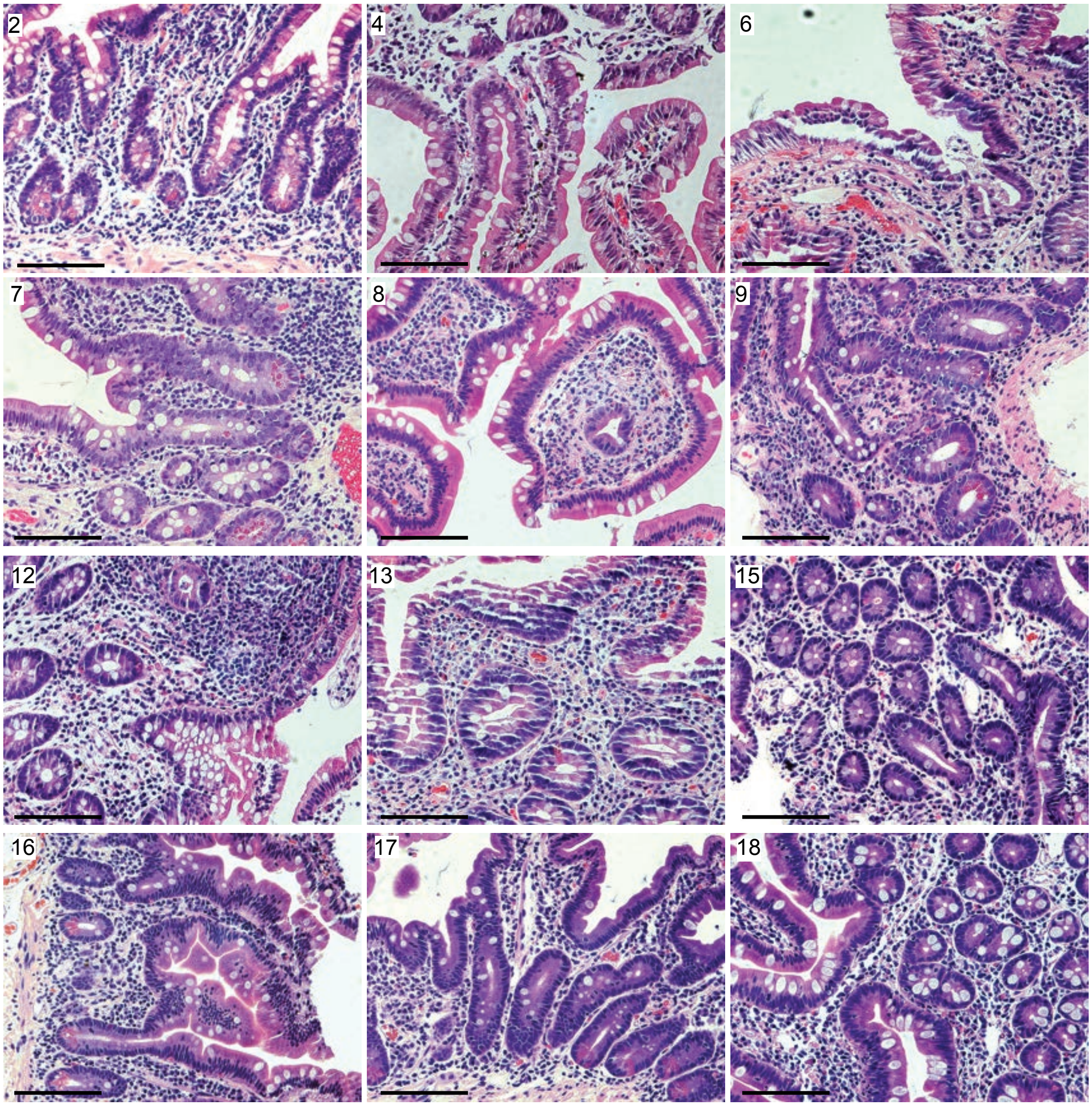

**Fig. S2 Representative H&E staining of small intestinal biopsies of COVID-19 patients.** Patient number in the top left corner corresponds with the patient number in table S4. All biopsies are duodenal with the exception of patient 12 which is from the terminal ileum. Scale bar; 100µm.

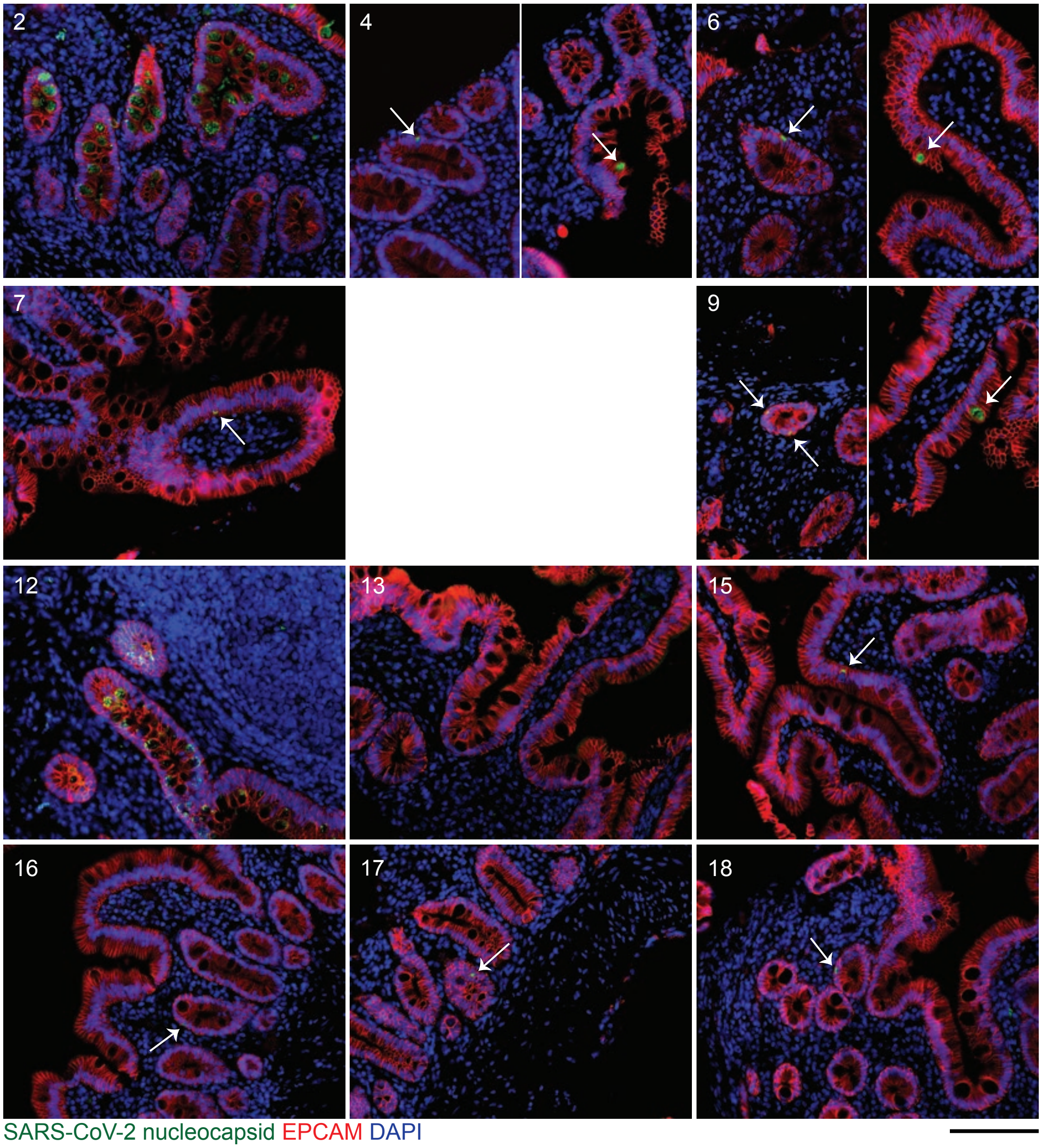

**Fig. S3 Representative immunofluorescence (IF) images of small intestinal biopsies of COVID-19 patients**. SARS-CoV-2 nucleocapsid (green), EPCAM (red) and DAPI (blue) in all COVID-19 patients where tissue was available for IF. Patient number in the top right corner corresponds with the patient number in table S4. All biopsies are duodenal with the exception of patient 12 which is from the terminal ileum. Patient 8 missing due to technical difficulties during IF staining. Scale bar; 100 µm.

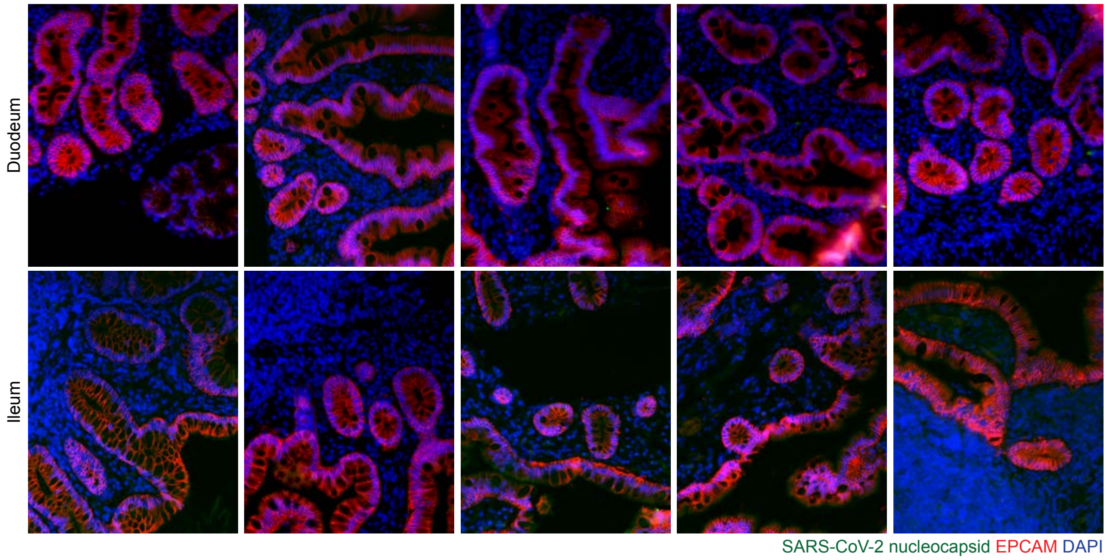

**Fig. S4 Representative immunofluorescence images of small intestinal biopsies of control patients.** SARS-CoV-2 nucleocapsid (green), EPCAM (red) and DAPI (blue) in duodenal biopsies (upper) and ileal biopsies (lower). Scale bar; 100 µm.

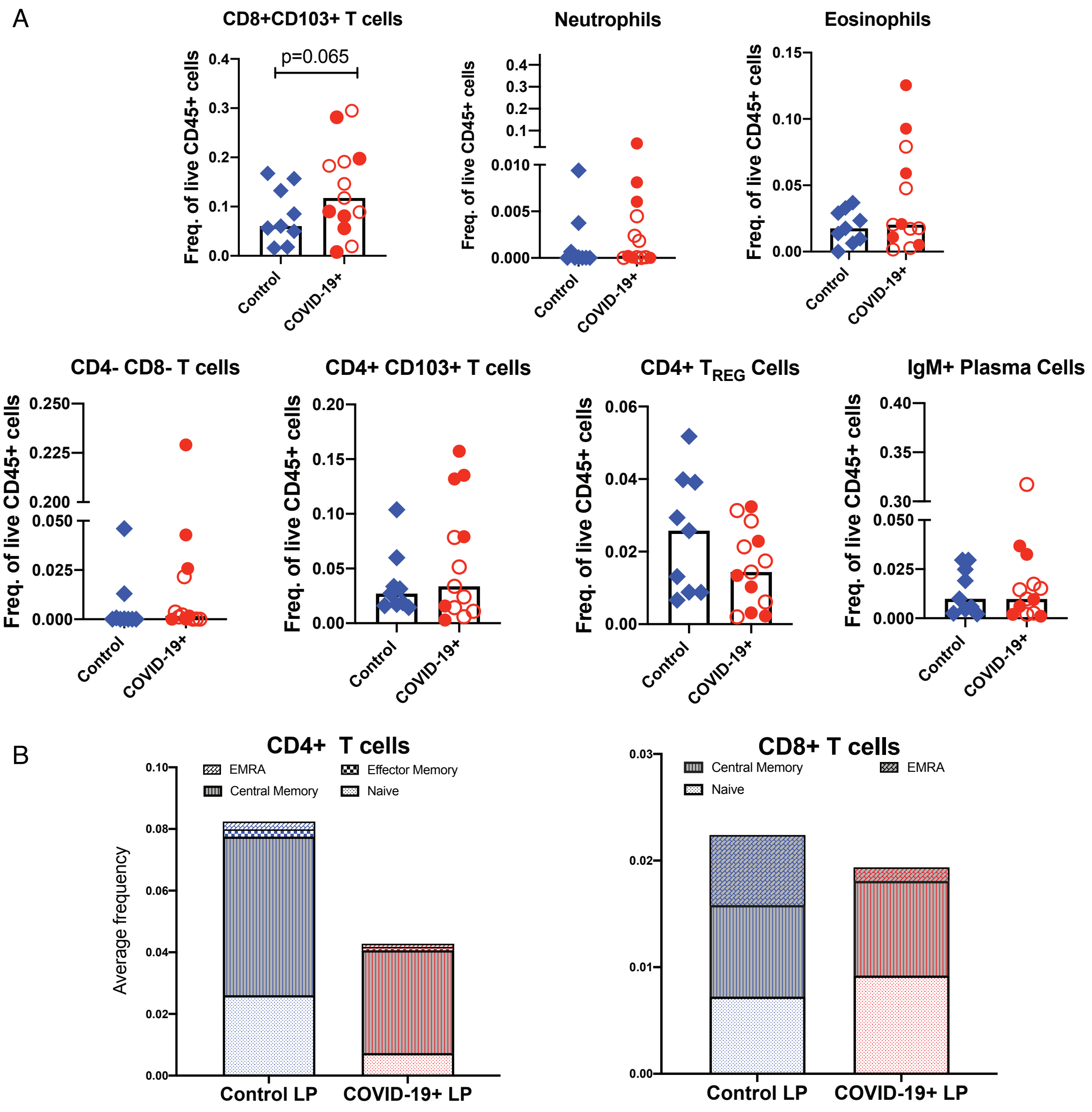

**Fig. S5 Altered immune populations in the lamina propria of COVID-19 patients compared to controls**. (**A**) Relative frequencies of lamina propria immune cells in controls and COVID-19 patients. Open red circles denote patients with asymptomatic/mild/moderate disease while filled red circles denote patients with severe COVID-19. The bar plots show median frequencies. (**B**) The stacked bar graphs show the distribution of average frequencies of naïve and memory CD4+ and CD8+ T cells in the lamina propria of COVID-19 patients and controls.

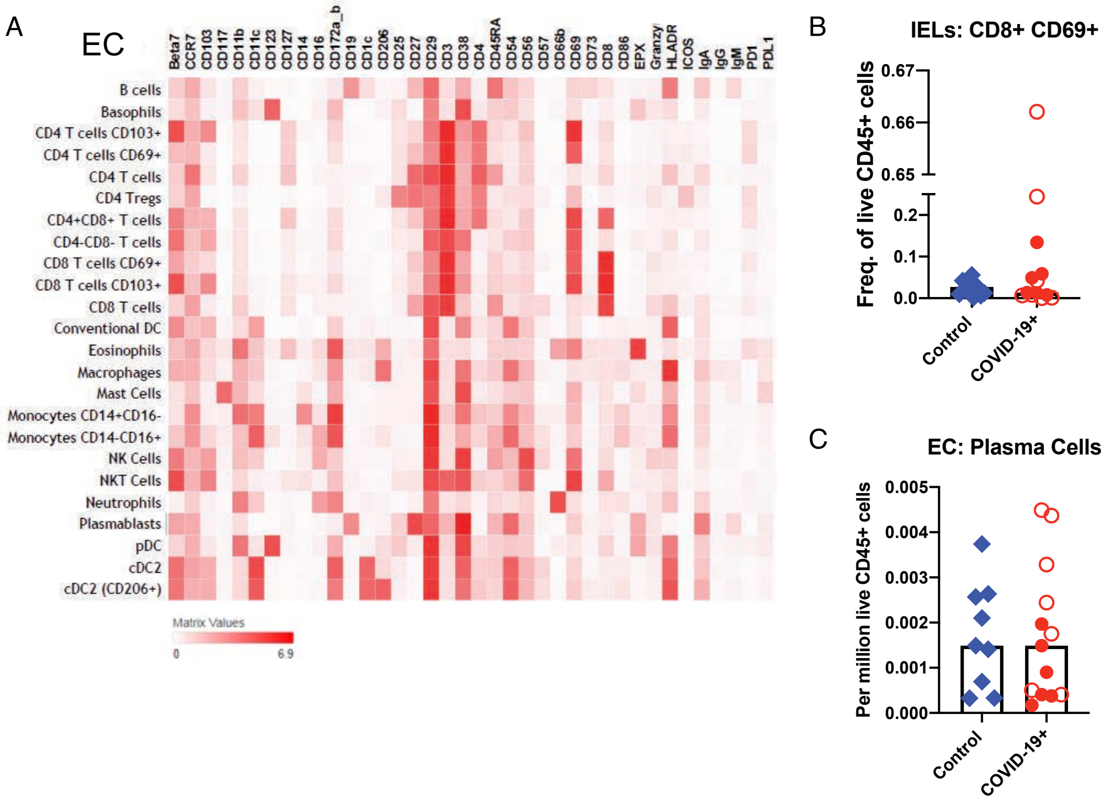

**Fig. S6 Altered immune populations in the epithelial compartment (EC) of COVID-19 patients compared to controls.** (**A**) The heat map shows clustering and distribution of different cell types in the EC. Relative frequencies of (**B**) intraepithelial lymphocytes (IELs) and (**C**) plasma cells in the EC of controls and COVID-19 patients. Open red circles denote patients with asymptomatic/mild/moderate disease while filled red circles denote patients with severe COVID-19. The bar plots show median frequencies.

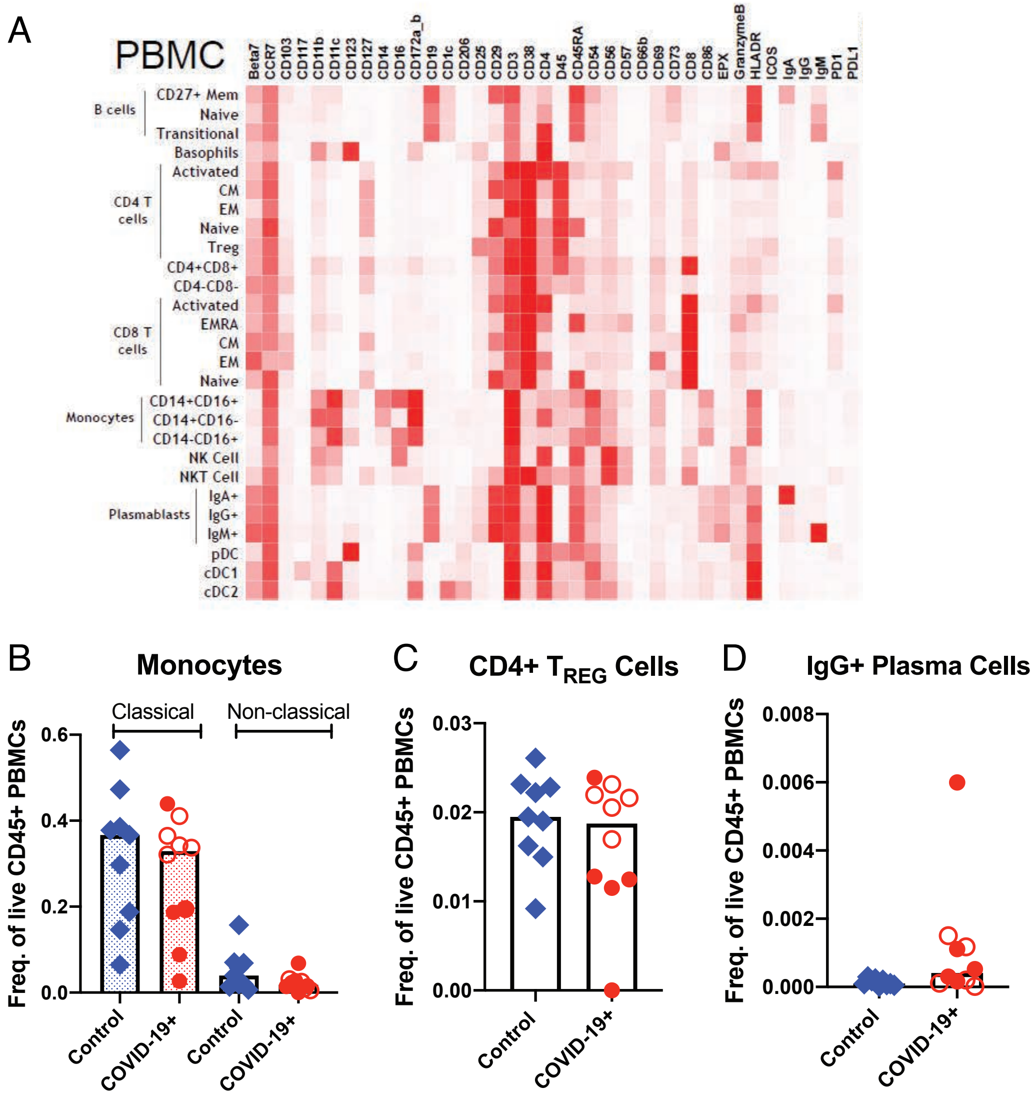

**Fig. S7 Altered immune populations in the blood of COVID-19 patients compared to controls.** (**A**) The heat map shows clustering and distribution of different immune cell types in the blood. Relative frequencies of (**B**) classical (dotted bars) and non-classical monocytes (open bars), (**C**) CD4+ regulatory T cells and (**D**) IgG+ plasma cells in the blood of controls and COVID-19 patients. Open red circles denote patients with asymptomatic/mild/moderate disease while filled red circles denote patients with severe COVID-19. The bar plots show median frequencies.

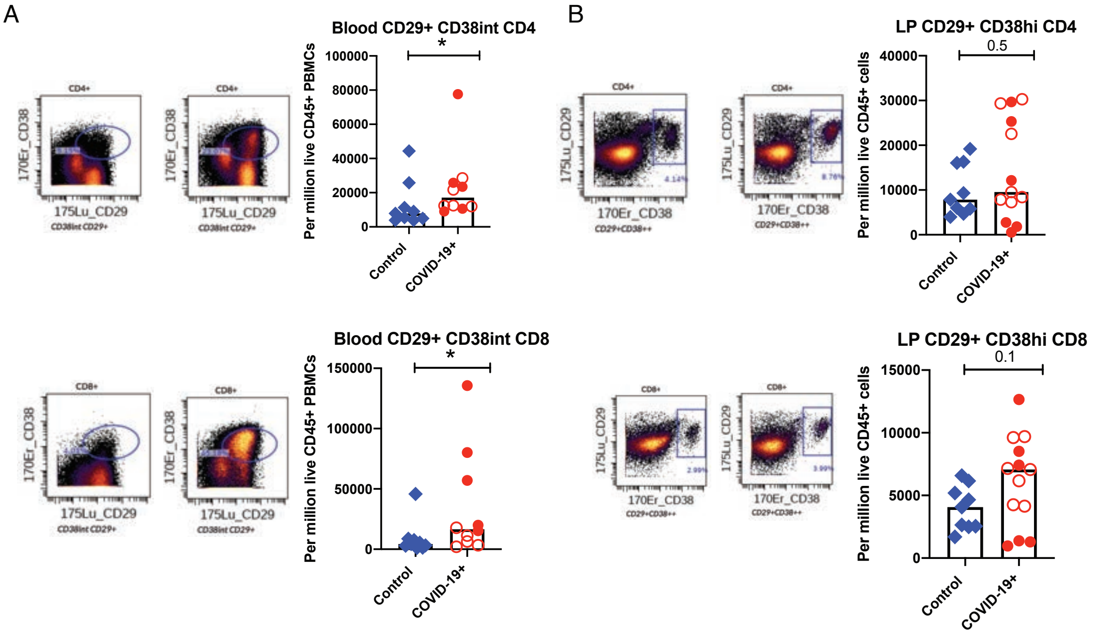

**Fig. S8 Altered T cell populations in blood and intestinal biopsies of COVID-19 patients compared to controls based on supervised analysis**. Representative CyTOF plots and bar plots comparing the frequencies of CD29+ CD38+ CD4+ and CD29+ CD38+ CD8+ T cells in (**A**) the blood and (**B**) lamina propria of controls (blue) and COVID-19 patients (red). Open red circles denote patients with asymptomatic/mild/moderate disease while filled red circles denote patients with severe COVID-19. The bar plots show median frequencies.

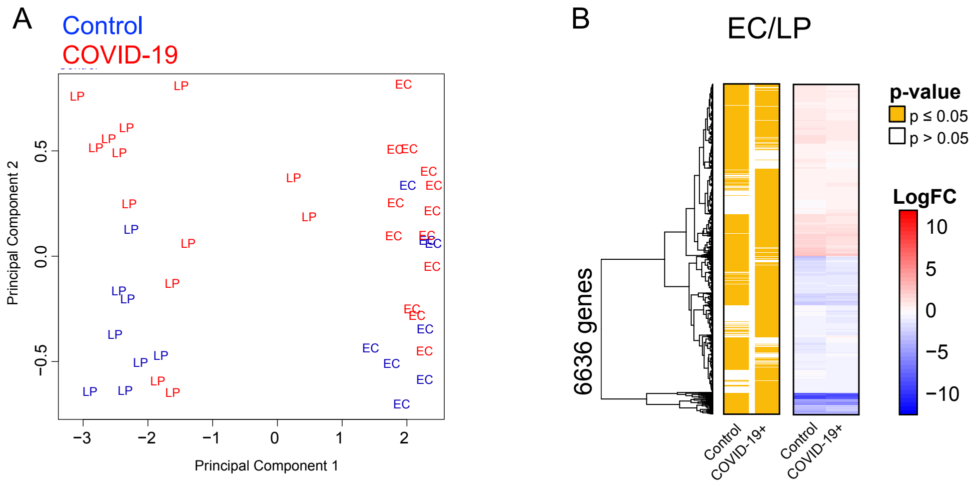

**Fig. S9 Distinct expression profiles in the intestinal epithelial compartment (EC) and lamina propria (LP).** (**A**) Principal Component Analysis (PCA) of EC and LP fractions of COVID-19 patients and controls. The two tissue fractions separate on the principal component 1 (x-axis). **(B)** Hierarchical clustering of average expression changes for 6636 genes (rows) characterizing the EC (red) or LP (blue) fractions (FDR ≤ 0.05) in the intestinal biopsies of COVID-19 patients and controls. The left panel indicates significant genes in yellow for each tissue compartment. The color bar (right panel) indicates the average log2 fold-change (FC).

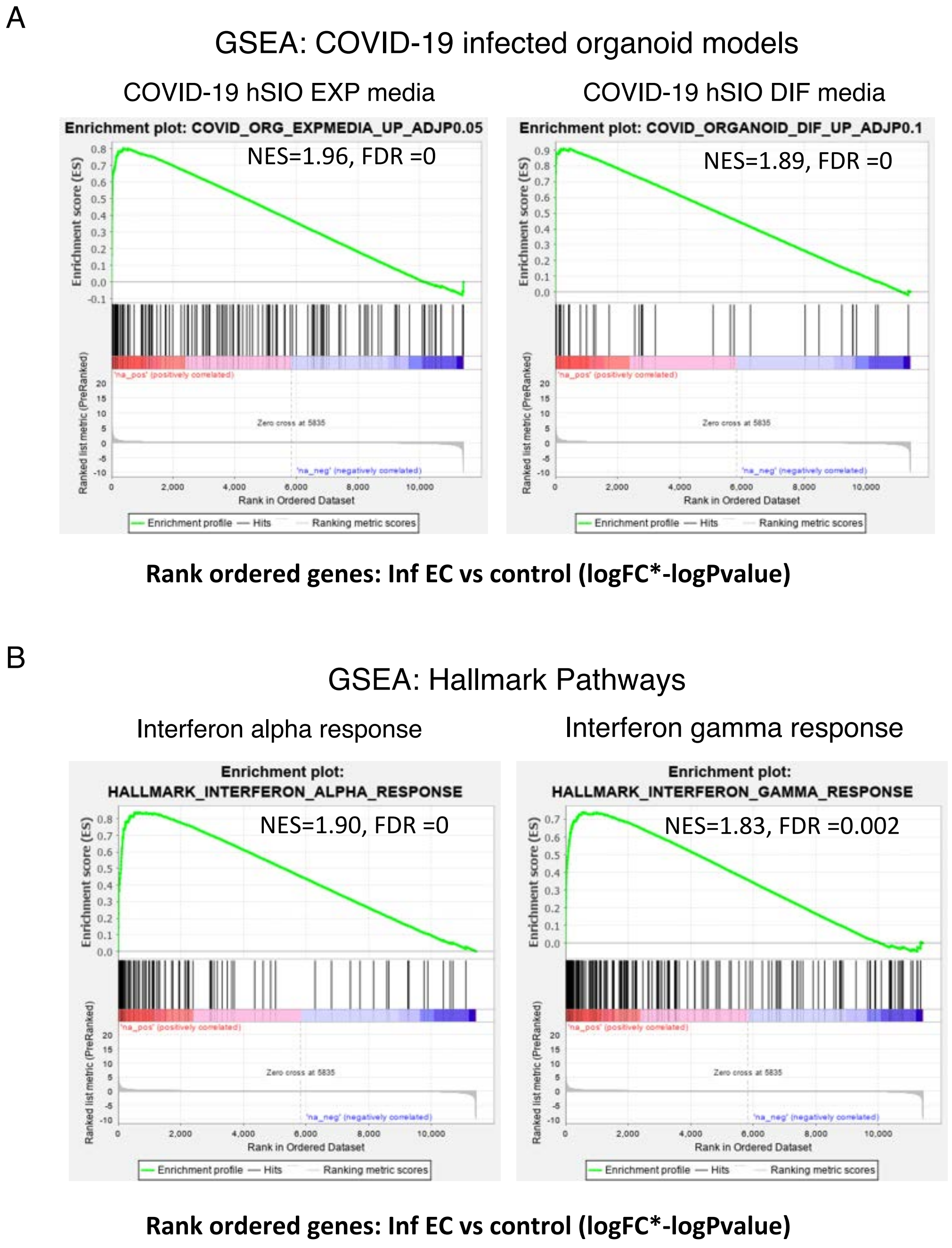

**Fig. S10 Immune signatures in the epithelial compartment (EC) of COVID-19 patients**. Gene Set Enrichment Analysis (GSEA) was performed using a rank ordered list of genes differentially expressed in the infected EC vs control EC. The metric for ranking was logFC*-logPvalue. (**A**) GSEA was performed on the rank ordered EC gene set using SARS-CoV-2 infected organoid datasets. The gene sets tested were molecular signatures curated from SARS-CoV-2 infected organoid experimental datasets using human small intestinal organoids (hSIOs) grown in either i) Wnt high expansion (EXP) medium (at adjP<0.05) or ii) differentiation (DIF) medium (at adjP<0.1). Only gene sets significantly enriched (at FDR <0.05) are displayed. (**B**) GSEA was performed for the same rank ordered EC gene set using the Hallmark Pathway datasets. Two significantly enriched pathways were found to be associated with upregulated genes in infected EC relative to controls (at FDR<0.05). Normalized enrichment score (NES) and FDR values are as indicated.

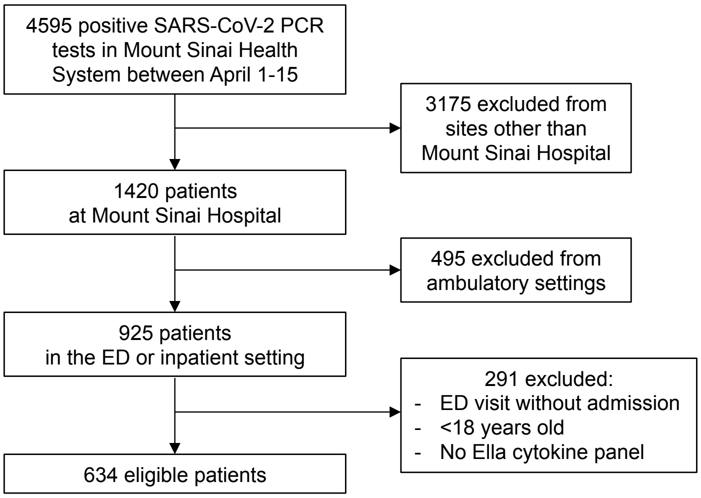

**Fig. S11 Flow diagram of the Discovery Cohort**. The diagram shows the total number of patients admitted to the Mount Sinai Health System between April 1-15, 2020 and the selection process that was adopted in order to select patients in the Discovery Cohort.

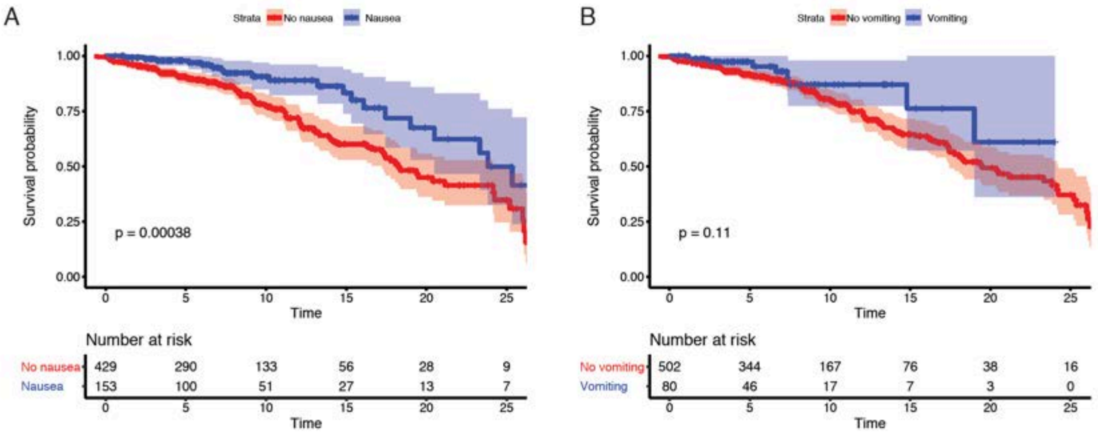

**Fig. S12 GI symptoms associated with reduced mortality and severity.** Kaplan-Meier curves for mortality stratified by (**A**) nausea and (**B**) vomiting for patients in the Discovery Cohort. P-values from log-rank test and 95% confidence intervals of Kaplan-Meier curves are shown. Below each Kaplan-Meier, the number of patients at risk for different time points are reported.

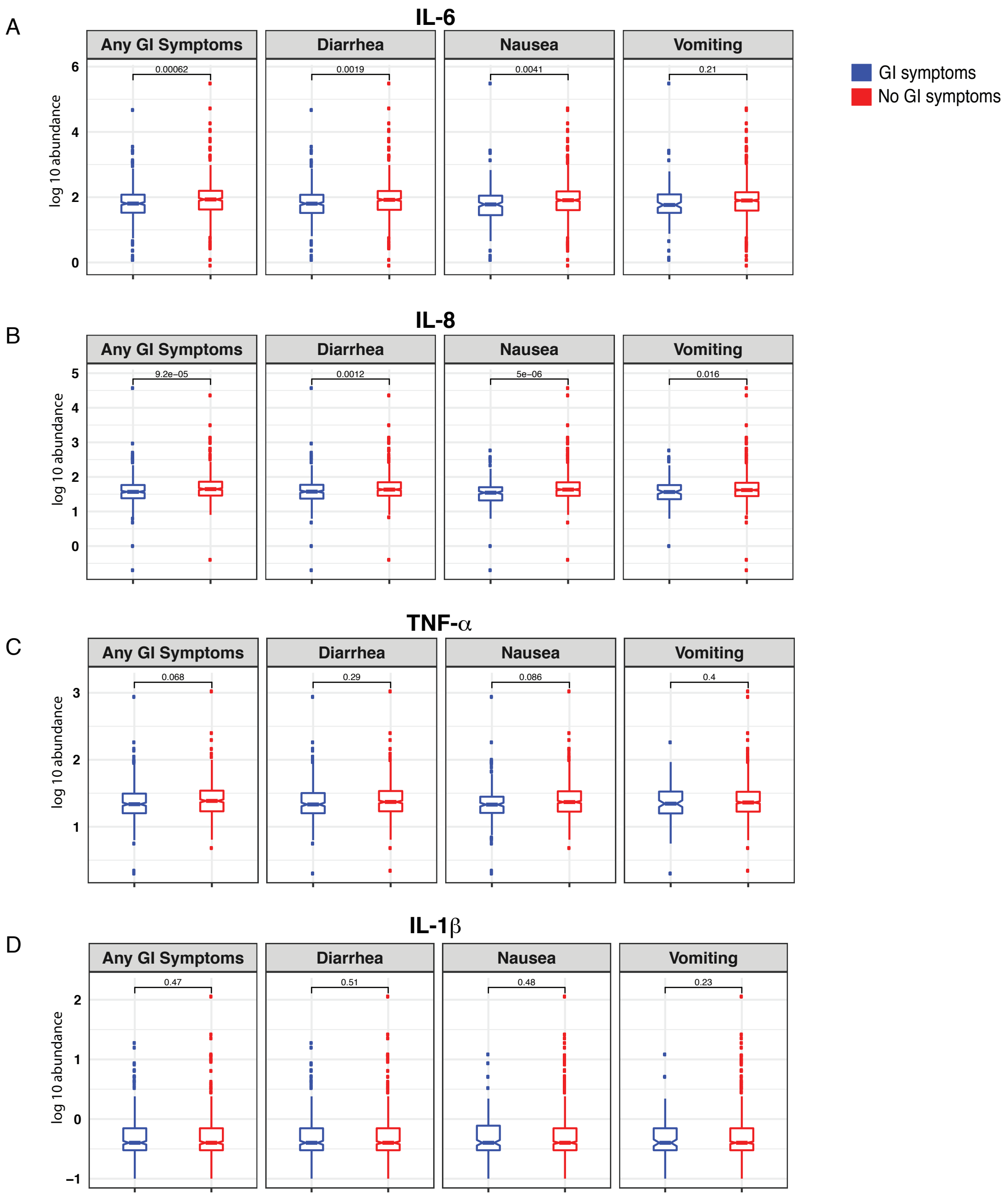

**Fig. S13 COVID-19 patients with GI symptoms have reduced levels of circulating IL-6 and IL-8**. (**A**) IL-6, (**B**) IL-8, (**C**) TNF-α and (**D**) IL-1β at the time of admission in patients with and without GI symptoms. Boxplots represent the median and interquartile range. P-values calculated using unpaired two-tailed t-test.

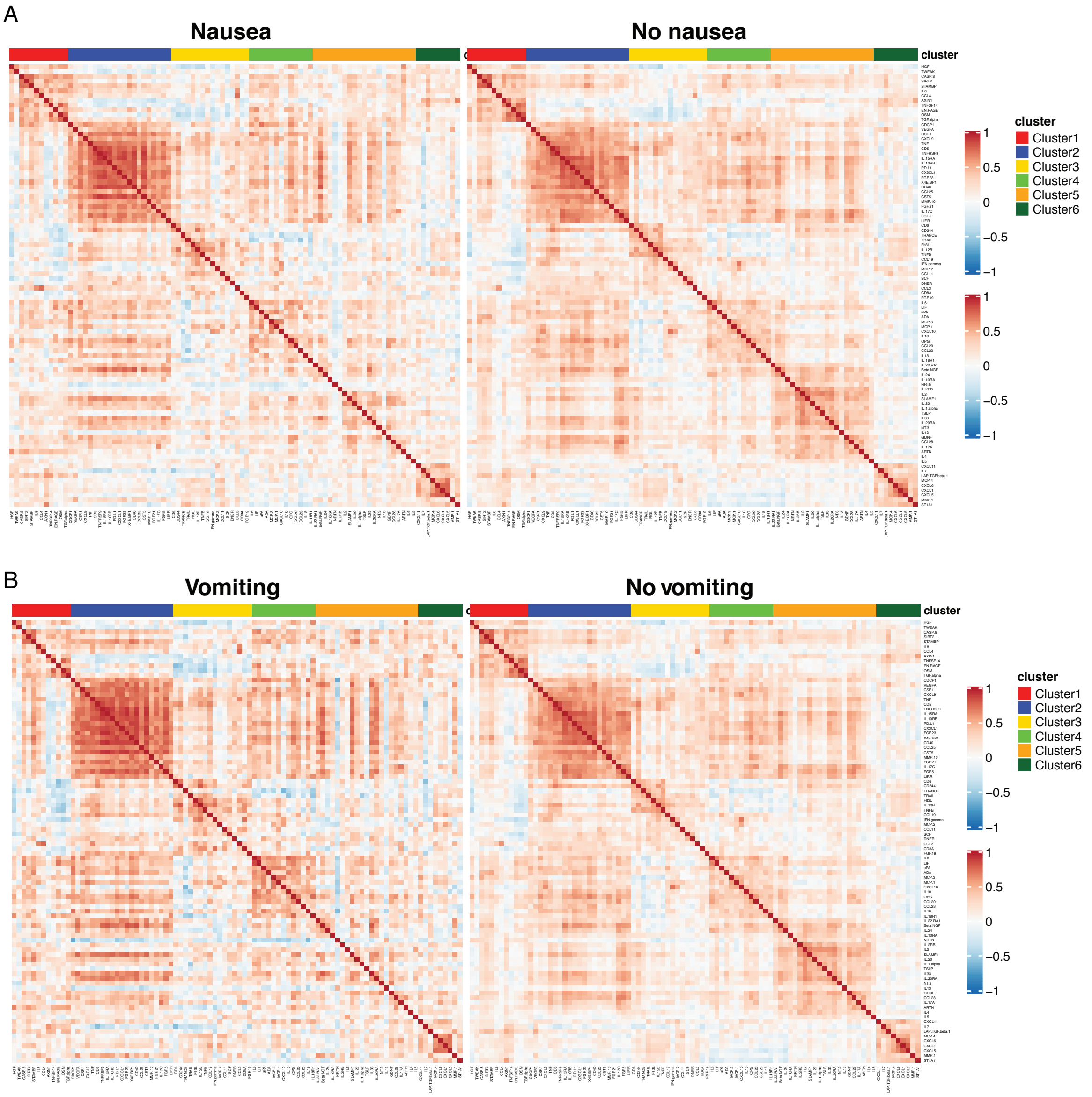

**Fig. S14 Correlation matrix (Pearson's) for 92 markers contained in the Olink platform.** (**A**) Correlation matrix across patients with nausea (left panel) compared to patients without nausea (right panel); and (**B**) patients with vomiting (left panel) compared to patients without vomiting (right panel). Cluster assignment derived using unsupervised consensus clustering is reported on the top of the heatmap.

**Table S1.** Clinical characteristics of SARS-CoV-2 uninfected control patients who underwent endoscopic biopsies of the GI tract.

| Control # | Age* | Sex | Days between negative NP SARS-CoV-2 swab and Procedure | Procedure | Tissue sample location |
|---|---|---|---|---|---|
| 1 | 70-75 | F | 1 | EGD | Duodenum |
| 2 | 60-65 | F | 1 | EGD | Duodenum |
| 3 | 65-70 | M | 2 | EGD & colonoscopy | Duodenum |
| 4 | 40-45 | M | 1 | EGD | Duodenum |
| 5 | 65-70 | M | 1 | EGD | Duodenum |
| 6 | 70-75 | M | 1 | EGD | Duodenum |
| 7 | 65-70 | F | 1 | EGD | Duodenum |
| 8 | 70-75 | F | 2 | EGD | Duodenum |
| 9 | 50-55 | M | 1 | EGD | Duodenum |
| 10 | 75-80 | F | 1 | Colonoscopy | Ileum |

* Age is provided as a range to obscure identifying information related to individuals
Abbreviations: EGD = Esophagogastroduodenoscopy, F = Female, M = Male

**Table S2.** Criteria for scoring disease severity in COVID-19 patients.

| **Severity Score** | **Criteria** |
|---|---|
| Mild | SpO2>94% on room air AND no pneumonia on imaging |
| Moderate | SpO2<94% on room air OR pneumonia on imaging |
| Severe | High flow nasal cannula (HFNC), non-rebreather mask (NRBM), Bilevel Positive Airway Pressure (non-invasive positive airway ventilation), or Mechanical ventilation AND no pressor medications AND creatinine clearance > 30 AND ALT < 5x upper limit of normal |
| Severe with end organ damage (EOD) | High flow nasal cannula (HFNC), non-rebreather mask (NRBM), Bilevel Positive Airway Pressure (non-invasive positive airway ventilation), or Mechanical ventilation AND pressor medications OR creatinine clearance <30 OR new renal replacement therapy OR ALT > 5x upper limit of normal |

**Table S3.** Clinical characteristics of COVID-19 patients including their symptoms on presentation, co-morbidities and treatment regimens.

| Patient # | Clinical setting | Symptoms on presentation | Other symptoms / findings / complications | Highest level of oxygen required | COVID-19 Treatments |
|---|---|---|---|---|---|
| 1 | Inpatient -ICU | CVA | Pneumonia on imaging | Mechanical ventilation | HCQ and azithromycin |
| 2 | Inpatient -ICU | Fever, shortness of breath, vomiting and diarrhea | | Room air | HCQ (given prior to admission at MSH) and steroids |
| 3 | Inpatient -non-ICU | Diarrhea and altered mental status | UTI, choledocholithiasis | Room air | Supportive care |
| 4 | Inpatient -ICU | Fever, cough, shortness of breath, abdominal pain | Cardiac arrest | Mechanical ventilation | HCQ, azithromycin, Remdesivir and therapeutic AC |
| 5 | Inpatient -non-ICU | Fever, nausea, vomiting and shortens of breath | | HFNC | Steroids and therapeutic AC |
| 6 | Outpatient | Asymptomatic at time of procedure, tested for procedure | Subsequently developed CVA | Room air | Therapeutic AC |
| 7 | Inpatient -non-ICU | Asymptomatic at time of procedure, tested for procedure | Dysphagia | Room air | Supportive care |
| 8 | Inpatient -ICU | Shortness of breath, altered mental status | UTI, bacteremia | HFNC | HCQ, steroids, therapeutic AC |
| 9 | Inpatient -ICU | Shortness of breath | | Mechanical ventilation | HCQ, azithromycin, steroids, therapeutic AC and convalescent plasma |
| 10 | NA | NA | NA | NA | NA |
| 11 | Inpatient -non-ICU | Shortness of breath, altered mental status | acute on chronic renal failure requiring HD | HFNC / BiPAP | HCQ, azithromycin, and steroids |
| 12 | Outpatient | Fatigue, myalgias, cough | | Room air | Supportive care |
| 13 | Outpatient | Chills, fever | Epigastric pain radiating to back | Room air | Supportive care |
| 14 | Inpatient -ICU | Shortness of breath, generalized weakness | | Mechanical ventilation | HCQ, azithromycin, steroids, therapeutic AC, convalescent plasma |
| 15 | Inpatient -non-ICU | Shortness of breath, abdominal pain | | Room air | Supportive care |
| 16 | Outpatient | Myalgias | Obstructive jaundice | Room air | Supportive care |
| 17 | Outpatient | Asymptomatic at time of procedure, tested for procedure | | Room air | Supportive care |
| 18 | Inpatient -non-ICU | Abdominal pain | Jaundice, UTI | Room air | Supportive care |

Abbreviations: CVA = cerebrovascular accident, HCQ = hydroxychloroquine, UTI = urinary tract infection, HFNC = high flow nasal cannula.

**Table S4.** Histopathological characteristics of COVID-19 patients.

| Patient # | Tissue used for IF | Viral protein detected by IF? | Increased IELs | Neutrophils | Other histopathologic abnormalities | Viral particles detected on EM? |
|---|---|---|---|---|---|---|
| 2 | Duodenum, ileum | yes (ileum and duodenum) | no | yes (ileum) | inflammation in ileum | not done |
| 3 | None available | N/A | yes (duodenum) | yes (mild, duodenum) | none | yes |
| 4 | Duodenum | yes | borderline | no | none | yes |
| 5 | None available | N/A | no | no | epithelial damage, villous blunting and reactive foveolar metaplasia | no |
| 6 | Duodenum | yes | no | yes | epithelial damage and foveolar metaplasia | no |
| 7 | Duodenum | yes | yes | yes (mild) | none | not done |
| 8 | None available | N/A | borderline | yes (mild) | none | no |
| 9 | Duodenum | yes | borderline | yes (mild) | epithelial damage | yes |
| 11 | None available | N/A | no | no | none | yes |
| 12 | Ileum | yes | no | no | none | yes |
| 13 | Duodenum | no | borderline | no | none | yes |
| 15 | Duodenum | yes | borderline | no | none | no |
| 16 | Duodenum | yes | no | no | none | no |
| 17 | Duodenum | yes | no | no | none | no |
| 18 | Duodenum | yes | patchy | yes (mild) | none | no |

Histologic information in reference to tissue used for IF unless not available in which case inflammatory changes as documented in EM report.

**Table S5.** Histopathological characteristics of pre-pandemic controls.

| Patient # | Age* | Sex | Tissue for IF | Reason for procedure | Medications |
|---|---|---|---|---|---|
| 1 | 50-55 | M | duodenum | epigastric pain | famotidine, ranitidine, azelastine-fluticasone, ipratropium bromide, levocetirizine, azelastine, omeprazole |
| 2 | 65-70 | F | duodenum and ileum | abdominal pain | none |
| 3 | 80-85 | F | duodenum | dysphagia | fluticasone-salmeterol, guaifenesin, albuterol, sennosides, Montelukast, losartan, amlodipine, acetaminophen, glipizide, atenolol, atorvastatin |
| 4 | 75-80 | F | duodenum | abdominal pain | losartan, aspirin, ferrous sulfate, metformin, omeprazole, pravastatin |
| 5 | 75-80 | F | duodenum | weight loss | spironolactone, apixaban, folic acid, torsemide, fluticasone |
| 6 | 30-35 | M | ileum | iron deficiency anemia | metformin, hydrochlorothiazide, enalapril |
| 7 | 55-60 | M | ileum | abdominal pain | divalproex, atorvastatin, benazepril, carvedilol, linagliptin, ondansetron, lamotrigine, lithium, rabeprazole, risperidone, alprazolam, oxycodone, glimepiride, allopurinol |
| 8 | 40-45 | M | ileum | rectal bleeding | none |
| 9 | 50-55 | F | ileum | CRC screening | acyclovir, acetaminophen |
| 10 | 20-25 | F | ileum | abdominal pain | none |

Abbreviation: CRC, Colorectal Cancer

**Table S6**. Discovery Cohort basic demographics, clinical characteristics and outcomes. For age, the mean ± standard deviation is listed. For categorical variables, the number of patients followed by the percent of patients in parentheses is listed.

| | **Discovery Cohort (n=634)** |
|---|---|
| Age (years) | 64.0 ± 15.7 |
| Male | 369 (58.2) |
| **Race/ethnicities** | |
| Hispanic | 177 (27.9) |
| African-American | 161 (25.4) |
| White | 137 (21.6) |
| Asian | 35 (5.5) |
| Other | 124 (19.6) |
| **Comorbidities** | |
| HTN | 229 (36.1) |
| Diabetes mellitus | 141 (22.2) |
| Obesity (BMI>30)* | 211 (37.1) |
| Chronic lung disease | 59 (9.3) |
| Heart disease | 111 (17.5) |
| Chronic kidney disease | 95 (15.0) |
| Cancer | 66 (10.4) |
| HIV | 11 (1.7) |
| IBD | 7 (1.1) |
| **Disease severity** | |
| Mild | 54 (8.5) |
| Moderate | 361 (56.9) |
| Severe | 158 (24.9) |
| Severe with EOD | 61 (9.6) |
| **Outcomes** | |
| ICU admission | 110 (17.4) |
| Mortality | 151 (23.8) |
| **GI symptoms** | |
| Nausea | 157 (24.8) |
| Vomiting | 82 (12.9) |
| Diarrhea | 245 (38.6) |
| Any GI symptoms | 299 (47.2) |

*BMI information available on 568/634 patients

**Table S7.** COVID-19 disease severity and mortality in patients with and without GI symptoms in the Discovery Cohort. Fisher's exact test used to calculate p-values.

| GI symptom | Severity | GI symptom | | p-value |
|---|---|---|---|---|
| | | Presence (n) | Absence (n) | |
| Nausea | Mild | 16 | 38 | 0.0112 |
| | Moderate | 102 | 259 | |
| | Severe | 32 | 126 | |
| | Severe EOD | 7 | 54 | |
| Vomiting | Mild | 10 | 44 | 0.0156 |
| | Moderate | 56 | 305 | |
| | Severe | 11 | 147 | |
| | Severe EOD | 5 | 56 | |
| Diarrhea | Mild | 26 | 28 | 0.0102 |
| | Moderate | 152 | 209 | |
| | Severe | 52 | 106 | |
| | Severe EOD | 15 | 46 | |
| Any GI symptoms | Mild | 31 | 23 | 0.0003 |
| | Moderate | 188 | 173 | |
| | Severe | 63 | 95 | |
| | Severe EOD | 17 | 44 | |

| GI symptom | Mortality | GI symptom | | p-value |
|---|---|---|---|---|
| | | Presence (n) | Absence (n) | |
| Nausea | Non-survivor | 21 | 130 | 0.0003 |
| | Survivor | 136 | 347 | |
| Vomiting | Non-survivor | 8 | 143 | 0.0008 |
| | Survivor | 74 | 409 | |
| Diarrhea | Non-survivor | 39 | 112 | 0.0002 |
| | Survivor | 206 | 277 | |
| Any GI symptoms | Non-survivor | 47 | 104 | <0.0001 |
| | Survivor | 252 | 231 | |

**Table S8.** Basic demographics in survivors and non-survivors in the Discovery Cohort. For age, the mean ± standard deviation and an unpaired two-tailed t-test was performed. For categorical variables, the number of patients followed by the percent of patients in parentheses is listed and the Fisher's exact test or the Chi-square test was used as appropriate.

| | **Survivors (n=483)** | **Non-survivors (n=151)** | **p-value** |
|---|---|---|---|
| Age (years) | 61.3 ± 15.2 | 72.6 ± 14.1 | **<0.0001** |
| Male | 287 (59.4) | 82 (54.3) | 0.30 |
| **Disease severity** | | | |
| Mild | 48 (9.9) | 6 (4.0) | |
| Moderate | 318 (65.8) | 43 (28.5) | |
| Severe | 95 (19.7) | 63 (41.7) | |
| Severe with EOD | 22 (4.6) | 39 (25.8) | **<0.0001** |

**Table S9.** Confidence intervals of odds ratio based on 1000 bootstrap iterations for severity, mortality and ICU admission in the Discovery Cohort.

| **Severity** | **2.5%** | **50%** | **97.5%** |
|---|---|---|---|
| (Intercept) | 0.015 | 0.071 | 0.320 |
| **Any GI Symptom** | 0.378 | 0.559 | 0.844 |
| baseline.GenderMale | 0.939 | 1.380 | 2.090 |
| baseline.Age | 1.004 | 1.016 | 1.031 |
| baseline.DIABETES | 0.600 | 0.995 | 1.729 |
| baseline.BMI | 1.009 | 1.039 | 1.069 |
| baseline.RACE_ETHNICITY_BLACK OR AFRICAN-AMERICAN | 0.293 | 0.503 | 0.876 |
| baseline.RACE_ETHNICITY_HISPANIC | 0.688 | 1.188 | 2.035 |
| baseline.RACE_ETHNICITY_OTHER | 0.771 | 1.286 | 2.245 |
| baseline.HTN | 0.636 | 0.990 | 1.543 |
| baseline.Lung.Disease | 0.272 | 0.562 | 1.063 |
| baseline.Heart.Disease | 0.673 | 1.095 | 1.842 |
| | **2.5%** | **50%** | **97.5%** |
| (Intercept) | 0.013 | 0.060 | 0.259 |
| **Diarrhea** | 0.433 | 0.653 | 0.978 |
| baseline.GenderMale | 0.963 | 1.413 | 2.124 |
| baseline.Age | 1.005 | 1.017 | 1.033 |
| baseline.DIABETES | 0.604 | 1.012 | 1.747 |
| baseline.BMI | 1.008 | 1.037 | 1.067 |
| baseline.RACE_ETHNICITY_BLACK OR AFRICAN-AMERICAN | 0.300 | 0.521 | 0.920 |
| baseline.RACE_ETHNICITY_HISPANIC | 0.707 | 1.215 | 2.098 |
| baseline.RACE_ETHNICITY_OTHER | 0.777 | 1.311 | 2.280 |
| baseline.HTN | 0.634 | 0.977 | 1.529 |
| baseline.Lung.Disease | 0.267 | 0.546 | 1.042 |
| baseline.Heart.Disease | 0.686 | 1.113 | 1.826 |
| | **2.5%** | **50%** | **97.5%** |
| (Intercept) | 0.013 | 0.058 | 0.271 |
| **Nausea** | 0.329 | 0.563 | 0.880 |
| baseline.GenderMale | 0.932 | 1.364 | 2.089 |
| baseline.Age | 1.005 | 1.018 | 1.032 |
| baseline.DIABETES | 0.609 | 1.015 | 1.738 |
| baseline.BMI | 1.007 | 1.036 | 1.065 |
| baseline.RACE_ETHNICITY_BLACK OR AFRICAN-AMERICAN | 0.322 | 0.543 | 0.946 |
| baseline.RACE_ETHNICITY_HISPANIC | 0.749 | 1.264 | 2.169 |
| baseline.RACE_ETHNICITY_OTHER | 0.816 | 1.340 | 2.338 |
| baseline.HTN | 0.615 | 0.950 | 1.477 |
| baseline.Lung.Disease | 0.248 | 0.516 | 0.980 |
| baseline.Heart.Disease | 0.676 | 1.091 | 1.830 |
| | **2.5%** | **50%** | **97.5%** |
| (Intercept) | 0.012 | 0.054 | 0.236 |
| **Vomiting** | 0.190 | 0.399 | 0.732 |
| baseline.GenderMale | 0.938 | 1.395 | 2.107 |
| baseline.Age | 1.006 | 1.018 | 1.032 |
| baseline.DIABETES | 0.615 | 1.032 | 1.744 |
| baseline.BMI | 1.006 | 1.036 | 1.065 |
| baseline.RACE_ETHNICITY_BLACK OR AFRICAN-AMERICAN | 0.330 | 0.558 | 0.962 |
| baseline.RACE_ETHNICITY_HISPANIC | 0.766 | 1.286 | 2.191 |
| baseline.RACE_ETHNICITY_OTHER | 0.802 | 1.337 | 2.307 |
| baseline.HTN | 0.610 | 0.950 | 1.511 |
| baseline.Lung.Disease | 0.261 | 0.526 | 0.981 |
| baseline.Heart.Disease | 0.703 | 1.123 | 1.886 |

| **Mortality** | **2.5%** | **50%** | **97.5%** |
|---|---|---|---|
| (Intercept) | 0.000 | 0.003 | 0.022 |
| **Any GI Symptom** | 0.335 | 0.544 | 0.861 |
| baseline.GenderMale | 0.679 | 1.049 | 1.703 |
| baseline.Age | 1.036 | 1.053 | 1.074 |
| baseline.DIABETES | 0.527 | 0.930 | 1.605 |
| baseline.BMI | 1.010 | 1.043 | 1.081 |
| baseline.RACE_ETHNICITY_BLACK OR AFRICAN-AMERICAN | 0.529 | 1.035 | 1.959 |
| baseline.RACE_ETHNICITY_HISPANIC | 0.867 | 1.597 | 2.899 |
| baseline.RACE_ETHNICITY_OTHER | 0.450 | 0.878 | 1.630 |
| baseline.HTN | 0.600 | 1.007 | 1.665 |
| baseline.Lung.Disease | 0.342 | 0.778 | 1.577 |
| baseline.Heart.Disease | 0.652 | 1.154 | 2.028 |
| | **2.5%** | **50%** | **97.5%** |
| (Intercept) | 0.000 | 0.003 | 0.017 |
| **Diarrhea** | 0.388 | 0.638 | 0.985 |
| baseline.GenderMale | 0.702 | 1.076 | 1.737 |
| baseline.Age | 1.038 | 1.055 | 1.076 |
| baseline.DIABETES | 0.531 | 0.945 | 1.608 |
| baseline.BMI | 1.007 | 1.041 | 1.079 |
| baseline.RACE_ETHNICITY_BLACK OR AFRICAN-AMERICAN | 0.562 | 1.071 | 2.020 |
| baseline.RACE_ETHNICITY_HISPANIC | 0.901 | 1.640 | 2.898 |
| baseline.RACE_ETHNICITY_OTHER | 0.468 | 0.900 | 1.709 |
| baseline.HTN | 0.595 | 1.001 | 1.665 |
| baseline.Lung.Disease | 0.333 | 0.752 | 1.561 |
| baseline.Heart.Disease | 0.660 | 1.166 | 2.020 |
| | **2.5%** | **50%** | **97.5%** |
| (Intercept) | 0.000 | 0.003 | 0.019 |
| Nausea | 0.255 | 0.490 | 0.886 |
| baseline.GenderMale | 0.669 | 1.041 | 1.719 |
| baseline.Age | 1.038 | 1.055 | 1.075 |
| baseline.DIABETES | 0.528 | 0.938 | 1.623 |
| baseline.BMI | 1.006 | 1.040 | 1.077 |
| baseline.RACE_ETHNICITY_BLACK OR AFRICAN-AMERICAN | 0.591 | 1.112 | 2.116 |
| baseline.RACE_ETHNICITY_HISPANIC | 0.956 | 1.718 | 3.085 |
| baseline.RACE_ETHNICITY_OTHER | 0.474 | 0.916 | 1.703 |
| baseline.HTN | 0.579 | 0.960 | 1.571 |
| baseline.Lung.Disease | 0.313 | 0.699 | 1.457 |
| baseline.Heart.Disease | 0.655 | 1.155 | 2.007 |
| | **2.5%** | **50%** | **97.5%** |
| (Intercept) | 0.000 | 0.002 | 0.016 |
| **Vomiting** | 0.116 | 0.364 | 0.753 |
| baseline.GenderMale | 0.690 | 1.070 | 1.759 |
| baseline.Age | 1.038 | 1.055 | 1.076 |
| baseline.DIABETES | 0.558 | 0.970 | 1.652 |
| baseline.BMI | 1.006 | 1.040 | 1.077 |
| baseline.RACE_ETHNICITY_BLACK OR AFRICAN-AMERICAN | 0.609 | 1.145 | 2.144 |
| baseline.RACE_ETHNICITY_HISPANIC | 1.004 | 1.746 | 3.086 |
| baseline.RACE_ETHNICITY_OTHER | 0.463 | 0.907 | 1.697 |
| baseline.HTN | 0.586 | 0.974 | 1.620 |
| baseline.Lung.Disease | 0.318 | 0.717 | 1.489 |
| baseline.Heart.Disease | 0.670 | 1.186 | 2.025 |

| **ICU admission** | **2.5%** | **50%** | **97.5%** |
|---|---|---|---|
| (Intercept) | 0.03 | 0.17 | 1.11 |
| **Any GI Symptom** | 0.46 | 0.75 | 1.19 |
| baseline.GenderMale | 1.23 | 2.00 | 3.26 |
| baseline.Age | 0.98 | 0.99 | 1.01 |
| baseline.DIABETES | 0.91 | 1.70 | 3.23 |
| baseline.BMI | 0.98 | 1.02 | 1.05 |
| baseline.RACE_ETHNICITY_BLACK OR AFRICAN-AMERICAN | 0.26 | 0.57 | 1.20 |
| baseline.RACE_ETHNICITY_HISPANIC | 0.79 | 1.50 | 2.95 |
| baseline.RACE_ETHNICITY_OTHER | 0.48 | 0.98 | 1.99 |
| baseline.HTN | 0.34 | 0.61 | 1.08 |
| baseline.Lung.Disease | 0.15 | 0.63 | 1.45 |
| baseline.Heart.Disease | 0.20 | 0.51 | 0.97 |
| | **2.5%** | **50%** | **97.5%** |
| (Intercept) | 0.02 | 0.16 | 1.10 |
| **Diarrhea** | 0.45 | 0.76 | 1.25 |
| baseline.GenderMale | 1.23 | 2.00 | 3.27 |
| baseline.Age | 0.98 | 0.99 | 1.01 |
| baseline.DIABETES | 0.89 | 1.70 | 3.25 |
| baseline.BMI | 0.98 | 1.02 | 1.05 |
| baseline.RACE_ETHNICITY_BLACK OR AFRICAN-AMERICAN | 0.26 | 0.56 | 1.19 |
| baseline.RACE_ETHNICITY_HISPANIC | 0.78 | 1.50 | 2.96 |
| baseline.RACE_ETHNICITY_OTHER | 0.48 | 0.98 | 1.98 |
| baseline.HTN | 0.34 | 0.61 | 1.09 |
| baseline.Lung.Disease | 0.15 | 0.62 | 1.44 |
| baseline.Heart.Disease | 0.20 | 0.52 | 0.99 |
| | **2.5%** | **50%** | **97.5%** |
| (Intercept) | 0.02 | 0.14 | 0.95 |
| **Nausea** | 0.45 | 0.85 | 1.42 |
| baseline.GenderMale | 1.24 | 2.00 | 3.33 |
| baseline.Age | 0.98 | 0.99 | 1.01 |
| baseline.DIABETES | 0.91 | 1.74 | 3.31 |
| baseline.BMI | 0.98 | 1.02 | 1.05 |
| baseline.RACE_ETHNICITY_BLACK OR AFRICAN-AMERICAN | 0.27 | 0.60 | 1.25 |
| baseline.RACE_ETHNICITY_HISPANIC | 0.81 | 1.54 | 3.00 |
| baseline.RACE_ETHNICITY_OTHER | 0.49 | 0.99 | 1.97 |
| baseline.HTN | 0.33 | 0.59 | 1.06 |
| baseline.Lung.Disease | 0.14 | 0.60 | 1.40 |
| baseline.Heart.Disease | 0.20 | 0.51 | 0.98 |
| | **2.5%** | **50%** | **97.5%** |
| (Intercept) | 0.02 | 0.15 | 0.98 |
| **Vomiting** | 0.16 | 0.53 | 1.08 |
| baseline.GenderMale | 1.21 | 1.98 | 3.26 |
| baseline.Age | 0.98 | 0.99 | 1.01 |
| baseline.DIABETES | 0.92 | 1.74 | 3.31 |
| baseline.BMI | 0.98 | 1.02 | 1.05 |
| baseline.RACE_ETHNICITY_BLACK OR AFRICAN-AMERICAN | 0.27 | 0.60 | 1.26 |
| baseline.RACE_ETHNICITY_HISPANIC | 0.82 | 1.55 | 3.01 |
| baseline.RACE_ETHNICITY_OTHER | 0.48 | 0.99 | 1.96 |
| baseline.HTN | 0.34 | 0.61 | 1.07 |
| baseline.Lung.Disease | 0.15 | 0.62 | 1.43 |
| baseline.Heart.Disease | 0.20 | 0.52 | 0.99 |

**Table S10.** Age, gender and mortality in an External Validation (Italian) Cohort stratified by presence or absence of diarrhea on admission. For age, the mean ± standard deviation and an unpaired two tailed t-test was performed. For categorical variables, the number of patients followed by the percent of patients in parentheses is listed and the Fisher's exact test or the Chi-square test was used as appropriate.

| | **Diarrhea on admission (n=80)** | **No diarrhea on admission (n=207)** | **p-value** |
|---|---|---|---|
| Age (years) | 60.6 ± 13.9 | 65.5 ± 13.3 | **0.006** |
| Male | 46 (57.5) | 149 (72.0) | **0.024** |
| ICU admission | 9 (11.3) | 43 (20.8) | 0.06 |
| Mortality | 8 (10.0) | 49 (23.7) | **0.008** |
| Death or ICU admission | 16 (20.0) | 83 (40.1) | **0.001** |

**Table S11.** Confidence interval of AUC (95%) based on 1000 bootstrap iterations for severity, mortality and ICU admission in the Internal Validation Cohort.

| **Severity** | **2.50%** | **50%** | **97.50%** |
|---|---|---|---|
| Age + BMI | 0.539 | 0.587 | 0.598 |
| Age + BMI + Nausea | 0.567 | 0.608 | 0.619 |
| Age + BMI + Vomiting | 0.558 | 0.607 | 0.618 |
| Age + BMI + Diarrhea | 0.574 | 0.630 | 0.651 |
| Age + BMI + Any GI symptoms | 0.605 | 0.640 | 0.651 |

| **Mortality** | **2.50%** | **50%** | **97.50%** |
|---|---|---|---|
| Age + BMI | 0.685 | 0.700 | 0.702 |
| Age + BMI + Nausea | 0.698 | 0.717 | 0.722 |
| Age + BMI + Vomiting | 0.702 | 0.719 | 0.724 |
| Age + BMI + Diarrhea | 0.697 | 0.718 | 0.726 |
| Age + BMI + Any GI symptoms | 0.708 | 0.727 | 0.736 |

| **ICU admission** | **2.50%** | **50%** | **97.50%** |
|---|---|---|---|
| Age + BMI | 0.534 | 0.560 | 0.599 |
| Age + BMI + Nausea | 0.496 | 0.523 | 0.650 |
| Age + BMI + Vomiting | 0.488 | 0.515 | 0.626 |
| Age + BMI + Diarrhea | 0.562 | 0.649 | 0.667 |
| Age + BMI + Any GI symptoms | 0.570 | 0.647 | 0.670 |

**Table S12.** IL-6, IL-8, TNF-α, and IL-1β concentrations on admission in patients with and without GI symptoms. Benjamini adjusted p-values (signed - log10 scale) from t-test are reported. Association passing a 10% FDR are highlighted in yellow.

| | **Nausea** | **Vomiting** | **Diarrhea** | **Any GI Symptoms** |
|---|---|---|---|---|
| IL-6 | -1.958 | -0.473 | -2.226 | -2.484 |
| IL-8 | -4.098 | -1.440 | -2.302 | -3.133 |
| TNF-α | -0.815 | -0.311 | -0.406 | -0.864 |
| IL-1β | -0.295 | -0.473 | -0.295 | -0.295 |

**Table S13.** Cluster assignment for each of the 92 Olink analytes.

| Marker | Cluster |
|---|---|
| IL8 | 1 |
| AXIN1 | 1 |
| OSM | 1 |
| CCL4 | 1 |
| TGF.alpha | 1 |
| TNFSF14 | 1 |
| HGF | 1 |
| SIRT2 | 1 |
| EN.RAGE | 1 |
| CASP.8 | 1 |
| TWEAK | 1 |
| STAMBP | 1 |
| VEGFA | 2 |
| CDCP1 | 2 |
| IL.17C | 2 |
| CXCL9 | 2 |
| CST5 | 2 |
| FGF.23 | 2 |
| FGF.5 | 2 |
| LIF.R | 2 |
| FGF.21 | 2 |
| IL.15RA | 2 |
| IL.10RB | 2 |
| PD.L1 | 2 |
| MMP.10 | 2 |
| TNF | 2 |
| CD5 | 2 |
| X4E.BP1 | 2 |
| CD40 | 2 |
| CCL25 | 2 |
| CX3CL1 | 2 |
| TNFRSF9 | 2 |
| CSF.1 | 2 |
| CD8A | 3 |
| CD244 | 3 |
| TRAIL | 3 |
| CD6 | 3 |
| SCF | 3 |
| CCL11 | 3 |
| CCL19 | 3 |
| TRANCE | 3 |
| IL.12B | 3 |
| CCL3 | 3 |
| Flt3L | 3 |
| DNER | 3 |
| IFN.gamma | 3 |
| FGF.19 | 3 |
| MCP.2 | 3 |
| TNFB | 3 |

| Marker | Cluster |
|---|---|
| MCP.3 | 4 |
| OPG | 4 |
| uPA | 4 |
| IL6 | 4 |
| MCP.1 | 4 |
| IL18 | 4 |
| IL.18R1 | 4 |
| IL10 | 4 |
| CCL23 | 4 |
| CXCL10 | 4 |
| LIF | 4 |
| CCL20 | 4 |
| ADA | 4 |
| GDNF | 5 |
| IL.17A | 5 |
| IL.20RA | 5 |
| IL.2RB | 5 |
| IL.1.alpha | 5 |
| IL2 | 5 |
| TSLP | 5 |
| SLAMF1 | 5 |
| IL.10RA | 5 |
| IL.22.RA1 | 5 |
| Beta.NGF | 5 |
| IL.24 | 5 |
| IL13 | 5 |
| ARTN | 5 |
| IL.20 | 5 |
| CCL28 | 5 |
| IL33 | 5 |
| IL4 | 5 |
| NRTN | 5 |
| NT.3 | 5 |
| IL5 | 5 |
| IL7 | 6 |
| LAP.TGF.beta.1 | 6 |
| CXCL11 | 6 |
| CXCL1 | 6 |
| MCP.4 | 6 |
| MMP.1 | 6 |
| CXCL5 | 6 |
| CXCL6 | 6 |
| ST1A1 | 6 |

**Table S14.** Olink analytes in patients with and without GI symptoms. P-values from t-test comparing patients with and without GI symptoms. Signed Benjamini-Hochberg adjusted p-value (-log10 scale) are reported.

| | **Any GI Symptoms** | **Nausea** | **Vomiting** | **Diarrhea** |
|---|---|---|---|---|
| IL8 | -1.315 | -0.654 | -1.345 | -1.562 |
| VEGFA | 0.006 | 0.014 | -0.356 | -0.027 |
| CD8A | -0.823 | -0.175 | 0.193 | -0.823 |
| MCP.3 | -0.524 | -1.437 | -0.422 | -0.254 |
| GDNF | -1.355 | -1.345 | -0.014 | -1.490 |
| CDCP1 | -0.449 | -0.309 | -0.175 | -0.524 |
| CD244 | -0.023 | 0.110 | 0.126 | -0.017 |
| IL7 | 1.345 | 0.175 | -0.123 | 1.345 |
| OPG | -2.183 | -1.209 | -0.014 | -2.183 |
| LAP.TGF.beta.1 | 0.356 | -0.006 | -0.626 | 0.407 |
| uPA | -0.156 | -0.009 | 0.126 | -0.385 |
| IL6 | -1.063 | -1.022 | -0.254 | -0.747 |
| IL.17C | -1.209 | -0.287 | -0.023 | -1.455 |
| MCP.1 | -0.175 | -0.654 | -0.458 | -0.187 |
| IL.17A | -2.183 | -1.097 | -1.419 | -2.183 |
| CXCL11 | 0.058 | -0.195 | -0.004 | 0.027 |
| AXIN1 | -0.009 | -0.031 | -1.209 | -0.026 |
| TRAIL | 1.063 | 0.626 | 0.314 | 0.458 |
| IL.20RA | -0.548 | -0.573 | -0.563 | -0.618 |
| CXCL9 | -0.573 | -0.367 | 0.023 | -1.209 |
| CST5 | -0.626 | -0.187 | -0.004 | -0.969 |
| IL.2RB | -0.156 | -0.028 | 0.009 | -0.044 |
| IL.1.alpha | 0.046 | -0.023 | 0.001 | 0.162 |
| OSM | -0.178 | -0.117 | -0.175 | -0.242 |
| IL2 | -0.341 | -0.424 | -0.287 | -0.264 |
| CXCL1 | -0.058 | -0.533 | -0.733 | -0.164 |
| TSLP | 0.009 | 0.001 | -0.031 | -0.009 |
| CCL4 | -0.287 | -0.022 | 0.022 | -1.355 |
| CD6 | -0.001 | 0.027 | 0.363 | 0.001 |
| SCF | -0.245 | -0.022 | -0.114 | -0.082 |
| IL18 | -0.254 | -0.068 | 0.332 | -0.434 |
| SLAMF1 | -0.618 | -0.707 | -0.190 | -0.708 |
| TGF.alpha | -1.087 | -0.626 | -0.675 | -1.345 |
| MCP.4 | 0.191 | -0.175 | -0.461 | 0.058 |
| CCL11 | -0.327 | -0.440 | -1.209 | -0.260 |
| TNFSF14 | 0.014 | -0.014 | -0.434 | -0.012 |
| FGF.23 | -0.556 | -0.058 | 0.036 | -1.209 |
| IL.10RA | 0.027 | 0.218 | -0.044 | -0.026 |
| FGF.5 | -0.823 | -0.164 | -0.027 | -1.365 |
| MMP.1 | 0.110 | 0.156 | -0.218 | 0.027 |
| LIF.R | -0.347 | -0.009 | 0.310 | -0.358 |
| FGF.21 | -0.495 | -0.003 | 0.056 | -0.880 |
| CCL19 | -0.044 | -0.227 | -0.156 | -0.175 |
| IL.15RA | -1.345 | -0.175 | 0.022 | -2.177 |
| IL.10RB | -1.355 | -0.389 | -0.079 | -2.183 |
| IL.22.RA1 | -0.073 | -0.022 | -0.009 | -0.254 |
| IL.18R1 | -0.208 | -0.054 | 0.175 | -0.156 |
| PD.L1 | -0.441 | -0.195 | 0.031 | -0.702 |

| | | | | |
|---|---|---|---|---|
| Beta.NGF | -0.573 | -0.458 | -0.022 | -0.495 |
| CXCL5 | -0.014 | -0.144 | -0.424 | -0.042 |
| TRANCE | 0.933 | 0.458 | 0.236 | 0.654 |
| HGF | -0.626 | -0.333 | -0.009 | -0.529 |
| IL.12B | 0.060 | 0.058 | -0.009 | 0.038 |
| IL.24 | -0.536 | -0.218 | 0.023 | -0.618 |
| IL13 | -0.007 | 0.075 | 0.068 | -0.126 |
| ARTN | -1.209 | -1.365 | -0.618 | -1.345 |
| MMP.10 | -1.562 | -0.967 | -0.156 | -2.073 |
| IL10 | -0.379 | -0.377 | 0.058 | -0.270 |
| TNF | -0.643 | -0.175 | -0.027 | -0.932 |
| CCL23 | -0.023 | -0.218 | 0.001 | 0.001 |
| CD5 | -0.823 | -0.357 | -0.036 | -0.933 |
| CCL3 | -0.933 | -0.553 | -0.576 | -1.490 |
| Flt3L | 0.031 | -0.009 | -0.377 | -0.009 |
| CXCL6 | -0.156 | -0.385 | -0.347 | -0.385 |
| CXCL10 | -0.001 | -0.441 | -0.012 | 0.031 |
| X4E.BP1 | -0.733 | -0.211 | 0.064 | -1.223 |
| IL.20 | -0.247 | -1.022 | -0.270 | -0.195 |
| SIRT2 | -0.123 | -0.164 | -0.332 | -0.162 |
| CCL28 | -2.183 | -2.183 | -0.737 | -2.183 |
| DNER | 0.175 | 0.317 | 0.332 | 0.202 |
| EN.RAGE | -0.357 | -0.193 | -0.166 | -0.526 |
| CD40 | -0.823 | -0.079 | 0.001 | -1.490 |
| IL33 | -0.236 | -0.164 | 0.023 | -0.287 |
| IFN.gamma | 0.218 | 0.044 | 0.175 | 0.270 |
| FGF.19 | -0.458 | -0.001 | -0.036 | -1.355 |
| IL4 | -0.020 | -0.175 | -0.027 | -0.009 |
| LIF | -1.345 | -0.526 | -0.573 | -1.355 |
| NRTN | 0.001 | -0.168 | -0.036 | -0.012 |
| MCP.2 | 0.319 | -0.327 | -0.036 | 1.231 |
| CASP.8 | -0.438 | -0.377 | -0.264 | -0.731 |
| CCL25 | -0.264 | 0.025 | -0.075 | -0.626 |
| CX3CL1 | -0.556 | -0.164 | -0.009 | -0.810 |
| TNFRSF9 | -1.345 | -0.175 | -0.012 | -2.029 |
| NT.3 | -0.012 | -0.363 | 0.009 | -0.122 |
| TWEAK | -0.014 | 0.014 | -0.113 | -0.012 |
| CCL20 | -0.737 | -0.576 | 0.187 | -0.823 |
| ST1A1 | 0.156 | -0.025 | -0.175 | 0.068 |
| STAMBP | -0.201 | -0.171 | -0.175 | -0.377 |
| IL5 | 0.156 | -0.009 | 0.156 | -0.009 |
| ADA | -0.259 | -0.270 | 0.175 | -0.434 |
| TNFB | 0.009 | -0.009 | -0.036 | 0.014 |
| CSF.1 | -0.458 | -0.028 | 0.012 | -0.450 |

**Table S15.** List of antibodies used for microscopy studies.

| Antigen | Clone | Vendor | Catalogue number | Host | Conjugation | Dilution |
|---|---|---|---|---|---|---|
| ACE2 | Polyclonal | Abcam | ab15348 | rabbit | Unconjugated | 1:1000 |
| EPCAM | SPM491 | GeneTex | GTX34693 | mouse | Unconjugated | 1:100 |
| SARS-CoV-2 nucleocapsid | Polyclonal | N/A* | N/A | rabbit | Unconjugated | 1:2000 |
| No known specificity (isotype control) | Polyclonal | Abcam | ab37415 | rabbit | Unconjugated | variable |
| Yeast GAL4 (isotype control) | 15-6E10A7 | Abcam | ab170190 | mouse | Unconjugated | variable |
| Mouse IgG H&L | Polyclonal | Abcam | ab150116 | goat | Alexa Fluor 594 | 1:1000 |
| Rabbit IgG H&L | Polyclonal | Abcam | ab150077 | goat | Alexa Fluor 488 | 1:1000 |
